# Machine Learning to Understand Genetic and Clinical Factors Associated with the Pulse Waveform Dicrotic Notch

**DOI:** 10.1101/2021.12.09.21267484

**Authors:** Jonathan W. Cunningham, Paolo Di Achille, Valerie N. Morrill, Lu-Chen Weng, Seung Hoan Choi, Shaan Khurshid, Victor Nauffal, James P Pirruccello, Scott D. Solomon, Puneet Batra, Jennifer E. Ho, Anthony A. Philippakis, Patrick T. Ellinor, Steven A. Lubitz

**Affiliations:** Cardiovascular Division, Brigham & Women’s Hospital, Boston, Massachusetts, USA; Cardiovascular Disease Initiative, The Broad Institute of MIT and Harvard, Cambridge, Massachusetts, USA; Data Sciences Platform, The Broad Institute of MIT and Harvard, Cambridge, Massachusetts, USA; Cardiovascular Research Center, Massachusetts General Hospital, Boston, Massachusetts, USA; Demoulas Center for Cardiac Arrhythmias, Massachusetts General Hospital, Boston, Massachusetts, USA; Division of Cardiology, Massachusetts General Hospital, Boston, Massachusetts, USA; CardioVascular Institute and Division of Cardiology, Department of Medicine, Beth Israel Deaconess Medical Center, Boston, MA

## Abstract

**Background:** Absence of a dicrotic notch on finger photoplethysmography (PPG) is an easily ascertainable and inexpensive trait that has been associated with age and prevalent cardiovascular disease (CVD). However, the trait exists along a continuum, and little is known about its genetic underpinnings or prognostic value for incident CVD.

**Methods:** In 169,787 participants in the UK Biobank, we identified absent dicrotic notch on PPG and created a novel continuous trait reflecting notch smoothness using machine learning. Next, we determined the heritability, genetic basis, polygenic risk, and clinical relations for the binary absent notch trait and the newly derived continuous notch smoothness trait.

**Results:** Heritability of the continuous notch smoothness trait was 7.5%, compared with 5.6% for the binary absent notch trait. A genome wide association study of notch smoothness identified 15 significant loci, implicating genes including *NT5C2* (P=1.2×10^−26^), *IGFBP3* (P=4.8×10^−18^), and *PHACTR1* (P=1.4×10^−13^), compared with 6 loci for the binary absent notch trait. Notch smoothness stratified risk of incident myocardial infarction or coronary artery disease, stroke, heart failure, and aortic stenosis. A polygenic risk score for notch smoothness was associated with incident CVD and all-cause death in UK Biobank participants without available PPG data.

**Conclusion:** We found that a machine learning derived continuous trait reflecting dicrotic notch smoothness on PPG was heritable and associated with genes involved in vascular stiffness. Greater notch smoothness was associated with greater risk of incident CVD. Raw digital phenotyping may identify individuals at risk for disease via specific genetic pathways.

## Introduction

Finger photoplethysmography (PPG), the measurement of pulse volume waveforms using infrared light, is an inexpensive, non-invasive, and scalable test similar to pulse oximetry that may provide insight into a patient’s cardiovascular physiology.^1,2^ Arterial stiffness index (ASI), the most commonly used PPG metric, is associated with elevated blood pressure and incident cardiovascular disease (CVD).^3,4^ Another PPG phenotype, absence of the dicrotic notch, has been recognized as a marker of age and prevalent coronary artery disease since the 1970s.^5^ However, the biological mechanisms driving absent notch, and its prognostic value for CVD events, are unknown. Examining the genetic basis of observed phenotypes like absent notch has been an effective strategy for identifying biological mechanisms underlying CVD.

We investigated absent notch on the PPG waveform in 169,787 participants in the UK Biobank. First, to maximize power, we produced a machine learning-based predictor of absent notch on raw PPG waveform that yielded a continuous trait representing the likelihood of notch absence. Waveforms with absent or small notches were assigned higher values of this notch smoothness trait, while waveforms with large notches were assigned lower values. Second, we performed a genome wide association study of this continuous trait to elucidate genetic mechanisms underlying notch smoothness. Third, we characterized associations of notch smoothness with incident cardiovascular disease.

## Methods

### Study Population

The UK Biobank is a population-based, prospective cohort study that enrolled 502,513 participants age 40-69 years between 2006-2010.^6^ Finger PPG was performed in 169,787 participants at the first visit and 30,999 participants at the imaging visit who underwent simultaneous cardiac MRI. PPG at the first visit was introduced later in the enrollment period (2009-10) and was not performed in Scotland. Analysis of UK Biobank data was performed under application 17488 and approved by the Mass General Brigham institutional review board. Informed consent was obtained from all participants by the UK Biobank.

### Ascertainment of PPG measurements and clinical factors

PPG was performed using the PulseTrace PCA2 device (CareFusion, San Diego, CA), which transmits infrared light at 940nm through the finger and generates a pulse waveform of the light absorption during the cardiac cycle, which is proportional to the volume of blood in the finger pulp. Data collection was performed for 10-15 seconds on the index finger of the left arm at rest. If the initial waveform was poor quality, the sensor was moved to a larger finger or the thumb.^7^ Arterial stiffness index (ASI) was calculated as standing height divided by the time from the peak to notch of the waveform, and reported in the UK Biobank Data Showcase (Field ID 21021). The PulseTrace PCA2 software was designed to interpret even small inflections in the downward diastolic slope of the waveform as a dicrotic notch. When the software was unable to detect any notch, the device displayed the output “stiff” and the UK Biobank reported absence of the notch (Field ID 4206). ^8^

Clinical cardiovascular phenotypes were defined using reports from medical history interviews, *International Classification of Diseases-Ninth and -Tenth Revision* codes, operation codes, and death registry records (**Supplemental Table 1**). Indexed left ventricular end diastolic and end systolic volumes, and left ventricular ejection fraction, were measured from cardiac magnetic resonance images using a validated deep learning algorithm as previously described. ^9,10^

### Creation of a continuous notch smoothness trait using machine learning

A continuous trait representing a spectrum of tracings with progressively smoother dicrotic notches was created from the raw PPG tracing using a convolutional neural network based on the ResNet architecture. The model was trained to predict the binary absent notch label provided by the UK Biobank from the 100-length vector raw PPG tracing. The notch smoothness trait reflected the likelihood of absent notch, and was derived for all participants with PPG data. To avoid overfitting, 5 models trained on 80% of the participants were used to calculate trait values for the remaining 20%. The distribution of notch smoothness and a sample of waveforms representing the spectrum notch smoothness are shown in **Figure 1**. In an independent set of PPG tracings from the imaging visit not used in model training, the model that generated the notch smoothness trait discriminated the binary absent notch label very accurately (area under receiver operator characteristic curve 0.997).

**Figure 1:**
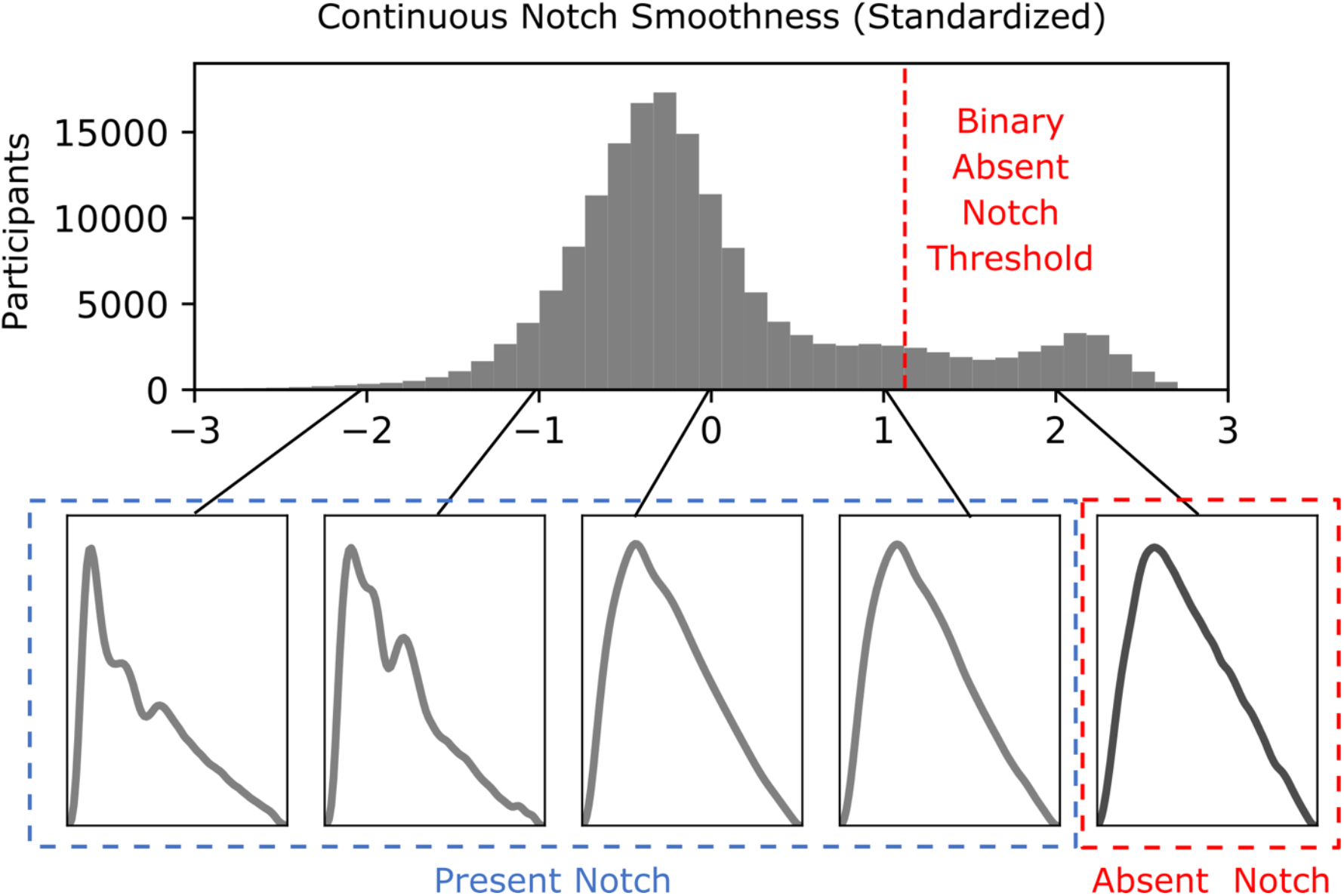
Transformation of Absent Notch to Continuous Notch Smoothness Using Supervised Machine Learning. The continuous notch smoothness trait reflects the likelihood of absent notch determined by the Resnet supervised machine learning model from interpretation of the raw PPG waveform. The distribution of the standardized notch smoothness is shown on top and sample PPG waveforms at each integer value are shown on bottom. The threshold for absence of a notch is illustrated in red.

### Genotyping and Genome-Wide Association Study

Participants in the UK Biobank were genotyped using the UK BiLEVE or UK Biobank Axiom arrays.^11^ Prior to imputation, variants with missingness >5%, minor allele frequency <0.00001, or violating Hardy-Weinberg equilibrium (P-value <1×10^−12^) were excluded. Samples with missingness >5%, excess heterozygosity, mismatch between reported and genotyped sex, or one of a pair of first- or second-degree relatives were excluded. Imputation was performed into the Haplotype Reference Consortium and UK10K+ 100G phase 3 reference panels using IMPUTE4. Variants with imputation INFO statistic <0.3 or on sex chromosomes were excluded. After exclusions, 7,843,256 variants were analyzed.

Genome wide association studies of absent notch and notch smoothness at the enrollment visit were performed using logistic and linear regression, respectively, assuming an additive genetic model and adjusted for age, sex, genotyping array and the first 12 principal components of ancestry using PLINK v2.00. Variants with P < 5 × 10^−8^ were considered significant. The lead SNP at each locus was determined by the smallest P-value, and independence of loci on the same chromosome were confirmed by conditional analysis. Test statistic inflation was estimated using LD score regression (ldsc v1.0.0).^12^ As a sensitivity analysis, association studies were repeated in only European ancestry participants, defined as self-identifying White participants after exclusion of outliers in 3 pairs of principal components (1-2, 3-4, and 5-6) using the R package *aberrant* (lambda=40), as previously described.^13,14^ Regional association plots were generated with LocusZoom using linkage disequilibrium data from the 1000G phase 3 European reference panel.^15^ SNP heritability was estimated using BOLT-REML v.2.3.4.^16^

A transcriptome-wide association study was performed to identify genes whose predicted expression was associated with notch smoothness, based on GWAS results. The S-PrediXcan package, Elastic Net, and eQTL data from GTEx version 8 were used.^17,18,19^ Target tissues of abdominal aorta and left ventricle were chosen based on the hypothesized physiology of notch smoothness. Since 7,571 genes were tested in aorta and 5,994 in left ventricle, Bonferroni-adjusted p-value threshold was set at 0.05/13,565 (7,571+5,994) = 3.7×10^−6^.

### Polygenic risk score

In a derivation cohort including 75% of participants with available PPG data, 20 candidate polygenic risk scores for notch smoothness were created from GWAS results by pruning and thresholding in Plink 1.9. Twenty permutations of p-value and linkage disequilibrium r^2^ thresholds were used: p-value thresholds 0.05, 5×10^−4^, 5×10^−6^, and 5×10^−8^; and r^2^ thresholds 0.05, 0.2, 0.4, 0.6 and 0.8. The top polygenic risk score (p-value<0.05 and r^2^<0.6; 183,888 variants) was selected based on the largest improvement in R^2^ (0.003) over the clinical model of age, sex, genotyping array, and the first 12 principal components of ancestry in a validation cohort of the remaining 25% of participants with available PPG data; univariate R^2^ was 0.002 (**Supplemental Table 2**). Patient-level polygenic risk score was then calculated using the top score in UK Biobank participants without available PPG data meeting the same genetic quality control criteria. Associations between standardized polygenic risk score and incident cardiovascular outcomes were assessed by multivariable Cox regression adjusted for age, sex, genotyping array, and the first 12 principal components of ancestry, excluding participants with prevalent cardiovascular disease or the outcome of interest.

### Association With Cardiovascular Diseases

Baseline characteristics of participants with and without absent notch and by quartile of notch smoothness at the enrollment visit were presented as number and proportion for categorical variables and mean and standard deviation for continuous variables. Prevalence of cardiovascular conditions at baseline was compared by multivariable logistic regression. Cardiovascular outcomes included hypertension, a composite of myocardial infarction or coronary artery disease (MI/CAD), heart failure, aortic stenosis, stroke, atrial fibrillation and all-cause death. The association of notch traits with incidence of these conditions or death was assessed by multivariable Cox regression, excluding participants with prevalent cardiovascular disease (defined as any of the above phenotypes except hypertension) at baseline. A composite of any cancer was included as a negative control. Three covariate models were considered. Model 1 adjusted for age at enrollment and sex alone. Model 2 added systolic blood pressure (SBP), diastolic blood pressure (DBP), heart rate, body mass index (BMI), prevalent diabetes, prevalent hypercholesterolemia, and ever smoking. Model 3 further adjusted for inverse rank-normalized arterial stiffness index measured by PPG, to evaluate whether notch morphology provided additive information to the most widely used PPG metric. We compared the risk stratification value of notch smoothness versus absent notch by calculating Harrell’s c-index for analogous models utilizing each trait as the primary exposure. Statistical analysis was performed using R version 4.1 (R Foundation for Statistical Computing, Vienna, Austria). Two-sided p-values <0.05 were considered significant.

We also examined the cross-sectional association between notch traits and cardiac MRI traits in patients who had both assessments. This analysis used PPG measurements from the imaging visit, which occurred after the enrollment visit. The association between notch traits and 3 cardiac MRI phenotypes—indexed left ventricular end diastolic and systolic volumes, and ejection fraction—was determined by linear regression with 3 levels of adjustment: unadjusted, adjusted for age and sex, and adjusted for age, sex, SBP, DBP, heart rate, and BMI.

## Results

### Baseline characteristics

Among the 169,787 participants who underwent PPG at the first visit, dicrotic notch was absent in 25,286 (14%) (**Supplemental Figure 1)**. Baseline characteristics of participants with and without a dicrotic notch are shown in **Supplemental Table 3**. Patients with absent notch were older (61.5 vs 56.5 years), more commonly female (66% vs 52%), and had a higher heart rate (70.1 vs 68.6 beats per minute), BMI (28.1 vs 27.4 kg/m^2^), and SBP (147.5 vs 139.3 mm Hg), but similar DBP. Prevalent cardiovascular diseases were rare but more common in patients with an absent notch, such as MI/CAD (6.4% vs 3.9%).

The continuous notch smoothness trait was associated with similar differences in baseline characteristics (**Table 1**). Most participants with binary absent notch (99.8%) had notch smoothness in the highest quartile. Notch smoothness was associated with other PPG traits: later position of the shoulder and peak of the waveform, greater arterial stiffness index, and greater wave reflection index (ratio of notch height to peak height). After adjustment for age, sex, heart rate, BMI, SBP, and DBP, smoking, diabetes mellitus, and hypercholesterolemia, greater notch smoothness was associated with greater prevalent cardiovascular conditions such as hypertension (adj. OR 1.07 [95% CI 1.06-1.08] per SD), MI/CAD (adj. OR 1.10 [95% CI 1.07-1.13] per SD, stroke (adj. OR 1.18 [95% CI 1.13-1.23] per SD), aortic stenosis (adj. OR 1.16 [95% CI 1.03-1.30] per SD) and atrial fibrillation (adj. OR 1.17 [95% CI 1.13-1.21] per SD, but not cancer (adj. OR 0.97 [95% CI 0.95-0.98]) (**Supplemental Table 4**).

**Table 1:** Baseline Characteristics by Quartile of Notch Smoothness.

|  | **Quartile 1** | **Quartile. 2** | **Quartile 3** | **Quartile 4** | **p-value** |
| --- | --- | --- | --- | --- | --- |
| n | 42447 | 42447 | 42447 | 42446 |  |
| Absent notch on PPG waveform | 2 (0.0) | 4 (0.0) | 41 (0.1) | 25239 (59.5) | <0.001 |
| Age, years | 55.3 (8.3) | 55.9 (8.1) | 57.1 (8.1) | 60.8 (6.8) | <0.001 |
| Male sex | 17870 (42.1) | 22571 (53.2) | 21508 (50.7) | 15812 (37.3) | <0.001 |
| Heart rate, beats per minute | 67.4 (10.8) | 69.3 (10.8) | 68.9 (11.3) | 69.7 (13.1) | <0.001 |
| Body mass index, kg/m<sup>2</sup> | 26.78 (4.63) | 27.55 (4.80) | 27.55 (4.83) | 27.95 (4.97) | <0.001 |
| Systolic blood pressure, mm Hg | 137.7 (19.5) | 138.0 (18.7) | 139.91 (19.3) | 146.26 (20.4) | <0.001 |
| Diastolic blood pressure, mm Hg | 81.7 (10.8) | 82.3 (10.6) | 82.2 (10.6) | 82.8 (10.8) | <0.001 |
| Ever smoking | 24166 (57.2) | 25055 (59.3) | 25273 (59.9) | 26186 (62.2) | <0.001 |
| Hypercholesterolemia | 5321 (12.5) | 6145 (14.5) | 6932 (16.3) | 9044 (21.3) | <0.001 |
| Diabetes mellitus | 937 (2.2) | 1117 (2.6) | 1333 (3.1) | 1806 (4.3) | <0.001 |
| Heart failure | 201 (0.5) | 232 (0.5) | 245 (0.6) | 327 (0.8) | <0.001 |
| Myocardial infarction or coronary artery disease | 1309 (3.1) | 1536 (3.6) | 1836 (4.3) | 2534 (6.0) | <0.001 |
| Aortic stenosis | 49 (0.1) | 54 (0.1) | 59 (0.1) | 118 (0.3) | <0.001 |
| Stroke | 471 (1.1) | 558 (1.3) | 608 (1.4) | 909 (2.1) | <0.001 |
| Atrial fibrillation | 539 (1.3) | 634 (1.5) | 726 (1.7) | 1056 (2.5) | <0.001 |
| Cancer | 3586 (8.4) | 3482 (8.2) | 3671 (8.6) | 4495 (10.6) | <0.001 |
| Position of shoulder, % of R-R cycle | 16.77 (5.23) | 20.48 (4.99) | 21.99 (4.61) | 25.32 (3.82) | <0.001 |
| Position of peak, % of R-R cycle | 18.77 (6.01) | 21.68 (4.97) | 22.89 (4.34) | 25.78 (3.51) | <0.001 |
| Position of notch, % of R-R cycle | 43.46 (6.52) | 43.13 (5.75) | 43.41 (6.04) | 44.23 (6.99) | <0.001 |
| Wave reflection index | 61.43 (27.74) | 67.72 (24.11) | 69.21 (27.01) | 72.53 (44.56) | <0.001 |
| Arterial stiffness index, m/s | 7.54 (2.64) | 9.22 (2.85) | 9.65 (4.50) | 10.94 (4.93) | <0.001 |

Values are expressed as mean (standard deviation) for continuous variables and number (percentage of total) for categorical variables.

### Genome wide association study of notch smoothness reveals 15 significant loci

We examined the SNP heritability of continuous notch smoothness compared with binary absent notch. Notch smoothness had a greater heritability at 7.5% (95% CI 6.8%-8.3%) versus 5.6% (95% CI 4.9%-6.4%) for the binary absent notch.

We then performed a genome wide association study of notch smoothness in 148,310 unrelated participants with PPG data, which revealed 15 loci at genome-wide significance (p < 5 × 10^−8^) (**Figure 2A, Table 2)**. Inflation (LD score regression intercept 1.013) was acceptable (**Supplemental Figure 2**). The strongest associations were identified at chromosome 10 near *NT5C2* and *CNNM2* (p=1.2×10^−26^), chromosome 7 near *IGFBP3* (p=4.8×10^−18^), chromosome 6 near *PHACTR1* (p=1.4×10^−13^), chromosome 14 near *SMOC1* (p=7.2×10^−12^), and chromosome 11 near *MYBPC3* (p=6.2×10^−10^) (**Supplemental Figure 3**). A similar GWAS using the binary absent notch phenotype identified only 6 genome-wide significant associations (**Figure 2B, Supplemental Table 5**), all at loci also identified for the continuous trait: chromosome 2 near *TEX41*, chromosome 3 near *ATP1B3*, chromosome 7 near *IGFBP3*, chromosome 10 near *NT5C2* and *CNNM2*, chromosome 11 near *MYBPC3*, and chromosome 14 near *SMOC1*. In a sensitivity analysis including only participants of European ancestry (n=123,644) 13 out of 15 loci significantly associated with notch smoothness remained associated (**Supplemental Table 6)**.

**Figure 2:**
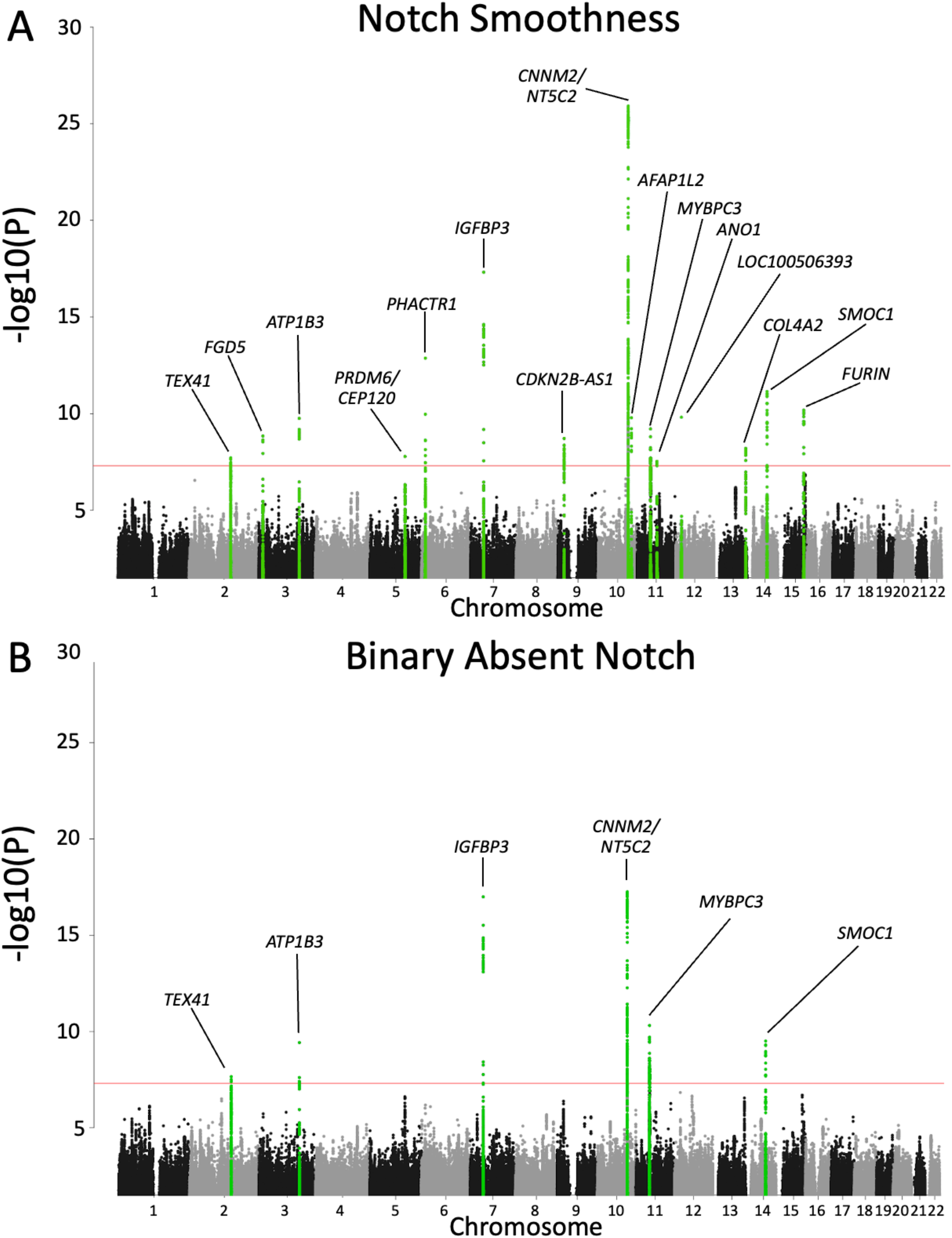
Genome Wide Association Study of Notch Smoothness and Binary Absent Notch. Genome-wide association study in 148,310 unrelated participants identifies 15 loci (green) associated with continuous notch smoothness (Panel A) and 6 loci associated with binary absent notch (Panel B). The significance threshold of a p-value less than 5×10^−8^ is indicated in red.

**Table 2:**
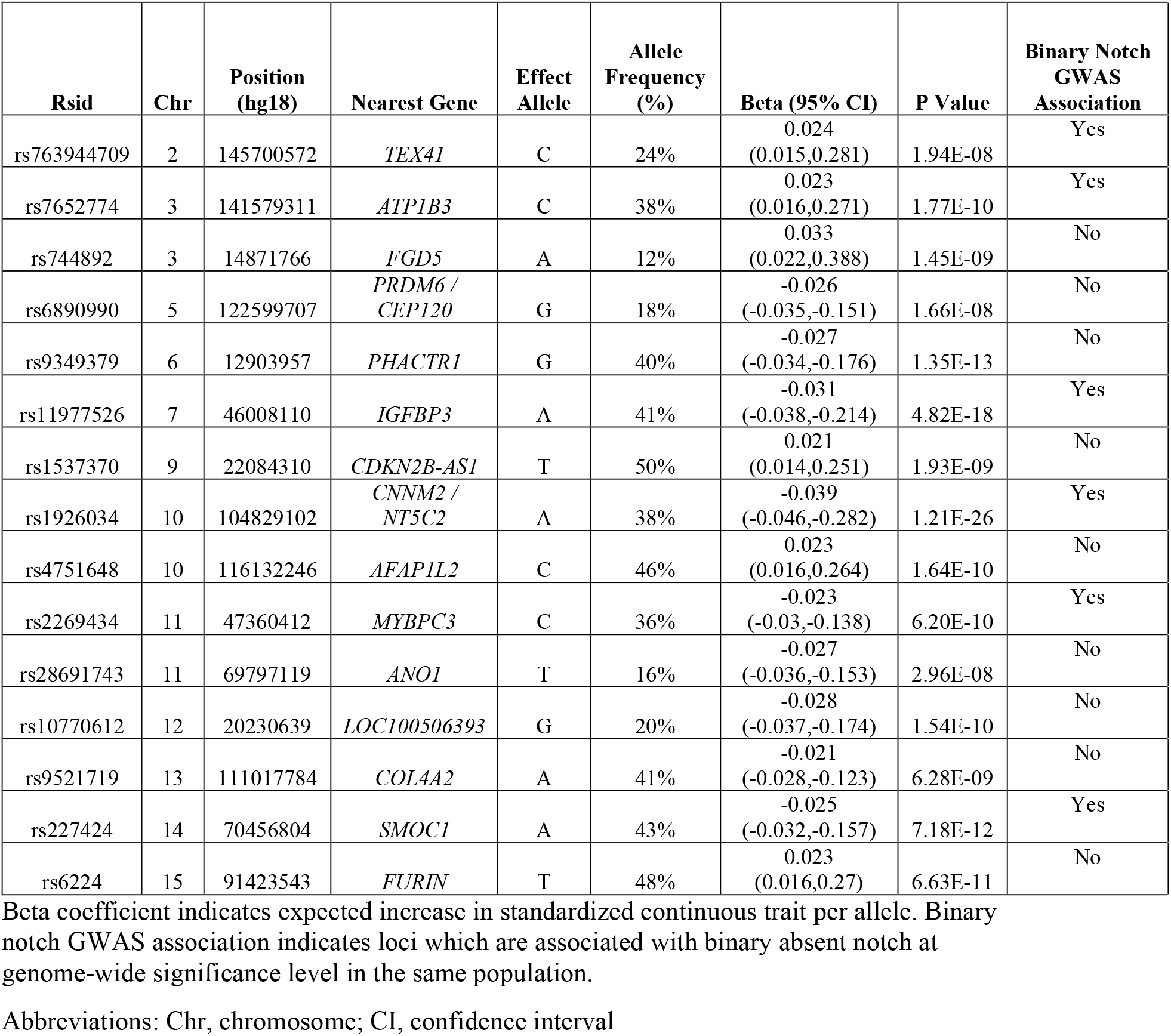
Variants Associated with Notch Smoothness at Genome-Wide Significance Threshold.

Finally, we performed a transcriptome wide association study to identify genes whose expression would be expected to vary significantly with notch smoothness (**Table 3**). We found 11 significant associations with genes in aortic tissue and 3 with genes in left ventricular tissue. The strongest associations were with *NT5C2, PHACTR1, SMOC1*, and *ATP1B3* in aortic tissue, and *IGFBP3, MFSD13A*, and *BORCS7* in left ventricle.

**Table 3:** Transcriptome Wide Association Study of Notch Smoothness.

| **Tissue** | **Gene** | **Standardized**<br>**Effect Size** | **Z score** | **P value** | **Variants**<br>**used** |
| --- | --- | --- | --- | --- | --- |
| Aorta | *NT5C2* | 0.40 | 9.2 | 3.7E-20 | 12 |
|  | *PHACTR1* | 0.16 | 6.9 | 5.7E-12 | 8 |
|  | *SMOC1* | -0.11 | -6.6 | 5.2E-11 | 10 |
|  | *ATP1B3* | 0.49 | 6.4 | 1.5E-10 | 6 |
|  | *AS3MT* | 0.03 | 6.2 | 5.1E-10 | 24 |
|  | *BORCS7* | 0.05 | 6.1 | 1.1E-09 | 24 |
|  | *ZEB2* | 0.28 | 5.7 | 1.1E-08 | 15 |
|  | *FES* | -0.13 | -5.3 | 1.0E-07 | 7 |
|  | *MFSD13A* | 0.05 | 5.1 | 3.3E-07 | 33 |
|  | *MRPS25* | 0.86 | 4.9 | 1.0E-06 | 1 |
|  | *PACSIN3* | -0.19 | -4.7 | 3.1E-06 | 24 |
| Left<br>Ventricle | *IGFBP3* | 0.31 | 8.1 | 6.3E-16 | 8 |
|  | *MFSD13A* | 0.09 | 7.2 | 5.9E-13 | 54 |
|  | *BORCS7* | 0.08 | 6.7 | 1.8E-11 | 17 |

Only associations with p value less than Bonferroni-corrected significance threshold are shown.

### Association of Notch Smoothness with Incident Cardiovascular Disease

Greater notch smoothness was associated with incident CVD and all-cause death in patients free of cardiovascular disease at the time of assessment (**Figure 3, Figure 4 & Supplemental Table 7)**. In a minimally adjusted model including age and sex, notch smoothness was associated with greater risk of hypertension, heart failure, MI/CAD, stroke, aortic stenosis and death, but not a negative control, incident cancer. All of these associations remained statistically significant after further adjustment BMI, heart rate, SBP, DBP, diabetes, hypercholesterolemia, and ever smoking. Risk discrimination of MI/CAD, heart failure, aortic stenosis, and stroke was stronger using notch smoothness compared to binary absent notch in univariable but not multivariable analyses (**Supplemental Tables 8 and 9**).

**Figure 3:**
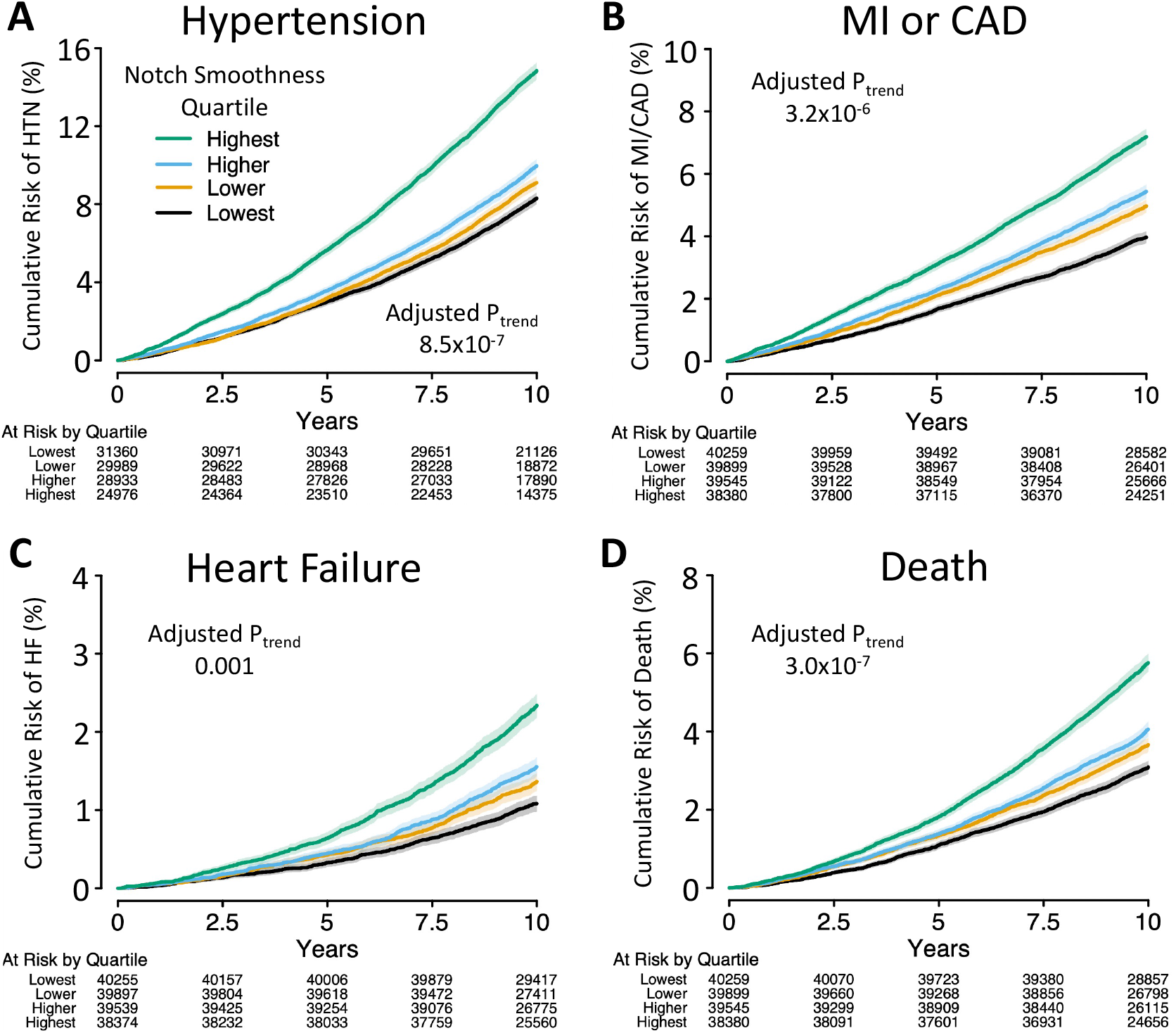
Cumulative Incidence of Cardiovascular Disease by Notch Smoothness Quartile. Higher notch smoothness quartile was associated with greater risk of incident hypertension, MI/CAD, heart failure, and all-cause death. Adjusted p-value for trend refers to a Cox proportional hazards regression model adjusted for age, sex, body mass index, heart rate, systolic and diastolic blood pressure, smoking, diabetes mellitus, and hypercholesterolemia. Participants with prevalent CVD (myocardial infarction or coronary artery disease, heart failure, stroke, aortic stenosis, or atrial fibrillation) at baseline were excluded. Participants with prevalent hypertension were excluded from analysis of incident hypertension only (Panel A).

**Figure 4:**
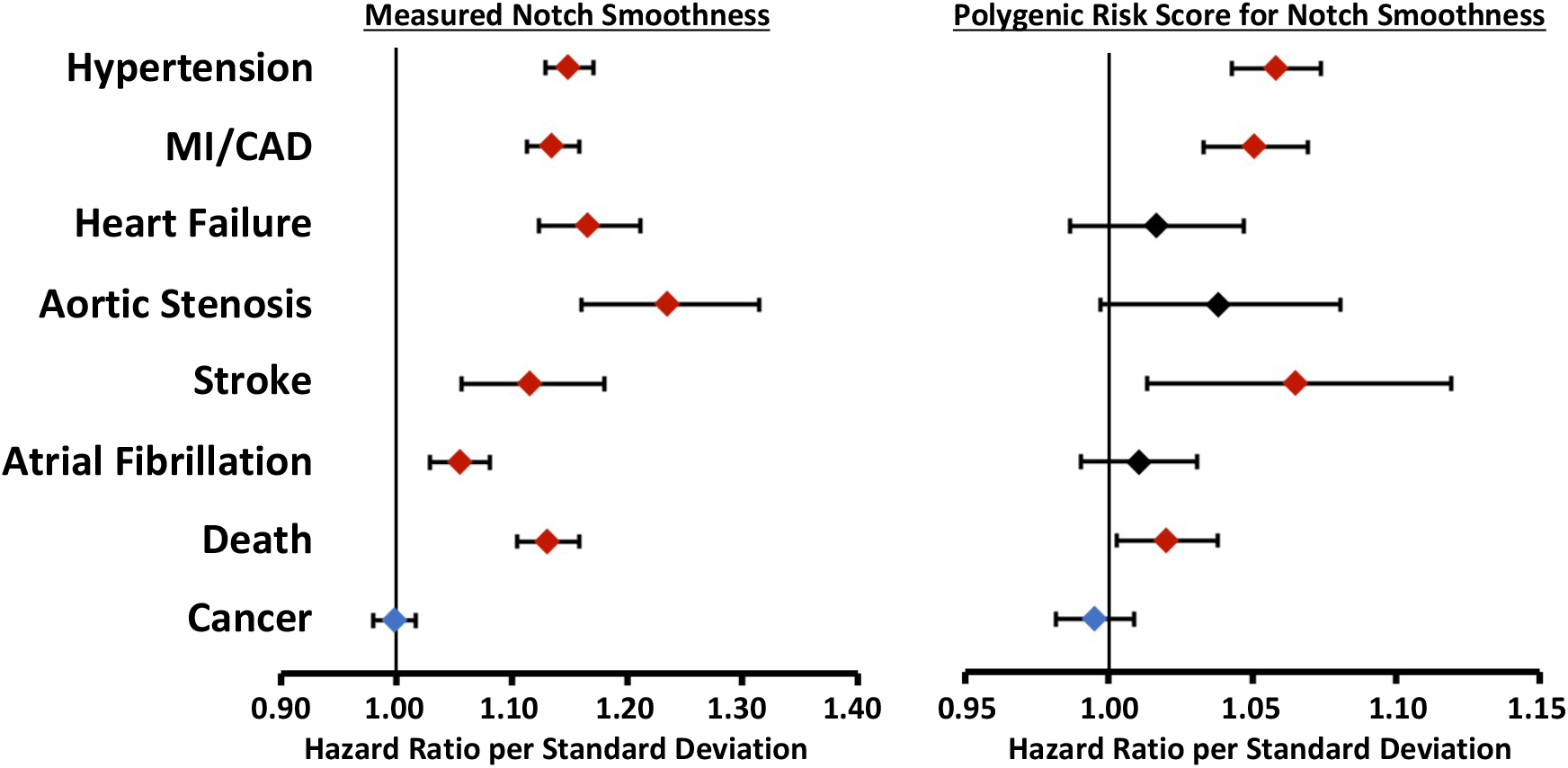
Prediction of Incident Cardiovascular Disease by Measured Notch Smoothness or Polygenic Risk Score for Notch Smoothness. Measured notch smoothness and polygenic risk score for notch smoothness predict incident cardiovascular disease, but not incident cancer. Left panel: Hazard ratio for incident disease per standard deviation of measured notch smoothness adjusted for age and sex, in participants with available PPG data. Right panel: Hazard ratio for incident disease per standard deviation of notch smoothness polygenic risk score, adjusted for age, sex, genotyping array, and the first 12 principal components of ancestry, in participants without available PPG data. Participants with prevalent CVD (myocardial infarction or coronary artery disease, heart failure, stroke, aortic stenosis, or atrial fibrillation) or the outcome of interest at baseline were excluded.

The prognostic value of notch smoothness was independent of, and superseded, the most commonly used PPG metric, arterial stiffness index (ASI), measured on the same PPG for all outcomes except hypertension (**Supplemental Table 7**). When notch smoothness and ASI were included in the same adjusted Cox regression, ASI was not associated with heart failure, aortic stenosis, or stroke while notch smoothness remained associated with these outcomes. However, ASI (HR 1.09 [95% CI 1.07-1.11] per SD, p=6.3×10^−16^) was more strongly associated with incident hypertension than notch smoothness (HR 1.03 [95% CI 1.01-1.05] per SD, p=0.01).

### Absent notch is associated with smaller left ventricular cavity size and higher left ventricular ejection fraction by cardiac MRI

PPG and cardiac MRI data were obtained simultaneously in 30,999 participants at the imaging visit. Participants with greater notch smoothness had smaller indexed left ventricular end diastolic volume (mean difference 1.8 ml/m^2^ [95% CI -1.7-2.0] per SD absent notch) and left ventricular end systolic volume (mean difference -1.1 ml/m^2^ [95% CI -1.0,-1.1] per SD) and higher ejection fraction (mean difference 0.6% [95% CI 0.5%, 0.7%] per SD) (all p<10^−28^). After adjustment for age, sex, SBP, DBP, heart rate, and BMI, notch smoothness remained associated with smaller end diastolic volume (mean difference -0.5 ml/m^2^ [95% CI -0.3,-0.6] per SD), end systolic volume (mean difference -0.4 ml/m^2^ [95% CI -0.3,-0.5] per SD) and higher ejection fraction (mean difference +0.3% [95% CI 0.3-0.4%] per SD) (all p<10^−12^). Similar associations were observed for binary absent notch (**Supplemental Table 10**).

### Polygenic risk score

The best polygenic risk score for the notch smoothness (linkage disequilibrium r^2^<0.6 and p<0.05; 183,888 variants) was selected based on most accurate prediction of observed notch smoothness in an independent validation cohort including 25% of participants with available PPG data (**Supplemental Table 2**). Among participants without PPG data, one standard deviation greater polygenic risk score for notch smoothness was associated with greater risk of incident hypertension (HR 1.06 [95% CI 1.04-1.07], p=2.5×10^−14^), MI/CAD (HR 1.05 [95% CI 1.03-1.07], p=1.8×10^−8^), stroke (HR 1.06 [95% CI 1.01-1.12], p=0.01) and death (HR 1.02 [95% CI 1.00-1.04], p=0.02) after adjustment for age, sex, genotyping array and the first 12 principal components and exclusion of participants with prevalent cardiovascular disease (**Figure 4**). Directionally consistent associations with incident heart failure and aortic stenosis did not reach statistical significance. The polygenic risk score was not significantly associated with the negative control outcome, incident cancer (HR 1.00 [95% CI 0.98-1.01], p=0.48).

## Discussion

Absent dicrotic notch on finger PPG is a marker of age and prevalent cardiovascular disease. In 169,787 UK Biobank participants, we derived a continuous trait reflecting a spectrum of dicrotic notch smoothness using machine learning interpretation of the raw pulse volume waveforms. A genome wide association of this continuous measure of dicrotic notch smoothness identified 15 loci, implicating genes including *NT5C2, ATP1B3*, and *IGFBP3* and pathways of vascular stiffness, compared with 6 loci for binary absent notch. A polygenic risk score for notch smoothness was associated with incident hypertension, coronary artery disease, stroke and death in UK Biobank participants without available PPG data. Notch smoothness was associated with incident cardiovascular disease independent of clinical risk factors and even arterial stiffness index measured on the same PPG. These results suggest that raw digital phenotyping may identify individuals at risk for disease through specific genetic pathways.

The primary innovation of this study is the creation of a novel continuous trait reflecting the dicrotic notch smoothness using machine learning. This trait represented our ResNet model’s prediction of the likelihood of absent notch based on the participant’s raw PPG waveform. For example, the model assigned low scores to participants with large dicrotic notches, intermediate scores to those with small but present notches, and high scores to those with absent notch. This notch smoothness trait was more heritable than binary absent notch, and a genome wide association study identified 15 rather genome wide significant associations compared with 6 for binary absent notch. These results illustrate that analysis of high-dimensional raw physiological data using machine learning can provide richer traits for genetic discovery.

Genome-wide and transcriptome wide association studies of notch smoothness identified loci known to be associated with cardiovascular disease, particularly hypertension (**Supplemental Table 11**). The strongest locus in our GWAS, chromosome 10 near *CNNM2 and NT5C2*, has been associated with systolic blood pressure. Knockdown of these 2 genes (but not other genes in the locus such as *AS3MT, BORCS7* or *CP17A1*) causes increased renin expression and arterial pulse in zebrafish. ^20,21^ *IGFBP3* inhibits IGF-1, which exerts pleiotropic effects including dilation of the peripheral vasculature through a nitric oxide dependent effect on vascular smooth muscle and endothelial cells.^22^ Higher circulating IGFBP3 is associated with greater mean arterial pressure. ^23^ *FURIN* is an upstream regulator of natriuretic peptides that is associated with blood pressure and coronary artery disease.^24,25^ *SMOC1* has been associated with hypertension, though with marginal significance in a large GWAS;^26^ our findings further nominate this gene in cardiovascular disease. *TEX41* has been associated with systolic blood pressure, aortic stenosis and arterial stiffness index by PPG. ^3,26,27^ Variants near *ATP1B3*, ^24^ *FGD5*, ^26,28^ *AFAP1L2*,^29^ and *MYBPC3*^28^ have also been associated with blood pressure traits, and *COL4A2* with coronary artery disease.^25^ Identification of *MYBPC3*, a top susceptibility gene for hypertrophic cardiomyopathy, raises the possibility that cardiac hypertrophy or dysfunction may affect dicrotic notch shape.

One genome-wide significant variant, rs9349379 near *PHACTR1*, offers a window into dicrotic notch physiology because the G allele is known to confer higher risk of coronary artery disease but lower risk of hypertension. In our study, the G allele was associated with lower values of notch smoothness, suggesting that hypertension rather than atherosclerosis is the primary driver of notch smoothness. This variant has been shown to affect vascular disease risk through expression of *EDN1*, which encodes the protein ET-1, a potent vasoconstrictor. ^30^

A polygenic risk score for notch smoothness was associated with cardiovascular diseases in UK Biobank participants without PPG data. This result indicates that inherited predisposition to smoother dicrotic notch modestly reflects predisposition to cardiovascular diseases. Future studies may examine whether raw digital health data such as PPG can define genetically mediated forms of disease with distinct physiology and potentially differential treatment response.

Smoother dicrotic notch by PPG is a meaningful cardiovascular trait associated with prevalent and incident disease independent of clinical risk factors and arterial stiffness index. In the 1970s, Dawber and colleagues demonstrated that absent notch was more common in older Framingham Heart Study participants with history of myocardial infarction.^5^ We extend these findings to incident CV events, independent of clinical risk factors, in a much larger cohort. The null association between notch smoothness and non-CV negative control, incident cancer, after minimal adjustment for age and sex suggests that notch smoothness is a specific marker of CV risk. While absent notch has been considered to reflect extreme arterial stiffness, the prognostic value of notch smoothness in our study was independent of arterial stiffness index for most clinical cardiovascular outcomes. PPG may be particularly useful in CV risk prediction because it is easily obtained, including from wearable devices.

These results must be interpreted in the context of the study design. Prevalent and incident cardiovascular disease events in the UK Biobank are derived from hospitalization records and self-report, and may be subject to misclassification. The UK Biobank is predominantly composed of individuals of European ancestry who were healthy at enrollment, and generalization of findings to other ancestral groups and individuals is unclear. The PPG signal was acquired over 15 seconds at a single encounter; averaged repeated measures may improve the precision of the phenotype. These results depend on the adjudication of binary absent notch, which was generated by the PulseTrace PCA2 software and provided by the UK Biobank.

## Conclusion

In 169,787 UK Biobank participants, we leveraged raw PPG data and supervised machine learning to develop a continuous trait reflecting smoothness of the dicrotic notch. A genome wide association of continuous notch smoothness identified 15 loci, many with known association with cardiovascular disease, particularly hypertension. A polygenic risk score for notch smoothness was associated with hypertension, coronary artery disease, stroke and all-cause death, suggesting that this phenotype shares underlying pathophysiology with multiple cardiovascular diseases. Measured notch smoothness was independently associated with risk of incident cardiovascular disease. These results provide a proof of concept that raw digital phenotyping may identify individuals at risk for disease through specific genetic pathways.

## Data Availability

This study involves only openly available human data, which can be obtained from the UK Biobank (https://www.ukbiobank.ac.uk/)

