## Supplemental Material for "Machine Learning to Understand Genetic and Clinical Factors Associated with the Pulse Waveform Dicrotic Notch"

**Table of Contents**

Supplemental Table 1: Clinical Outcome Definitions…………………………………………….2

Supplemental Table 2: Comparative Assessment of 20 Polygenic Risk Scores for Notch Smoothness ……………………………………………………………..……………10

Supplemental Table 3: Baseline Characteristics of Participants With and Without Binary Absent Notch…………………………..………………………..……..…………………11

Supplemental Table 4: Multivariable Adjusted Association of Binary Absent Notch & Notch Smoothness with Prevalent Cardiovascular Disease at Baseline …….…….…………..………..13

Supplemental Table 5: Variants Associated with Binary Absent Notch…......………..……..…14

Supplemental Table 6: Variants Associated with Notch Smoothness in Participants of European Ancestry ………………………………„„„„„„„„„…………………………...……...…..15

Supplemental Table 7: Association of Notch Smoothness with Incident Cardiovascular Events in Participants Free of Cardiovascular Disease at Baseline …………………...……...16

Supplemental Table 8: Association of Binary Absent Notch with Incident Cardiovascular Events in Participants Free of Cardiovascular Disease at Baseline …………..…17

Supplemental Table 9: C-Statistics for Incident Cardiovascular Disease With Binary Absent Notch and Notch Smoothness Traits..……………………………...………....18

Supplemental Table 10: Cross-Sectional Association of Notch Smoothness & Cardiac MRI Traits at Imaging Visit ……………….……………………..……………….19

Supplemental Table 11: Annotation of Genome Wide Significant Loci for Notch Smoothness……………………………………….……………………………………21

Supplemental Figure 1: CONSORT Diagram……………………………………………...23

Supplemental Figure 2: Q-Q Plot for Notch Smoothness GWAS ….…………..…….……...24

Supplemental Figure 3: Locus Zoom Plots for Loci Associated with Notch Smoothness...…...25

**Supplemental Table 1: Clinical Outcome Definitions**

Myocardial Infarction or Coronary Artery Disease

| Source | Code |
|---|---|
| ICD9 | 410 Acute myocardial infarction |
| ICD9 | 4109 Acute myocardial infarction |
| ICD9 | 411 Other acute and subacute forms of ischaemic heart disease |
| ICD9 | 4119 Other acute and subacute forms of ischaemic heart disease |
| ICD9 | 412 Old myocardial infarction |
| ICD9 | 4129 Old myocardial infarction |
| ICD9 | 4140 Coronary atherosclerosis |
| ICD9 | 4148 Other specified forms of chronic ischaemic heart disease |
| ICD9 | 4149 Chronic ischaemic heart disease, unspecified |
| ICD10 | I21 Acute myocardial infarction |
| ICD10 | I21.0 Acute transmural myocardial infarction of anterior wall |
| ICD10 | I21.1 Acute transmural myocardial infarction of inferior wall |
| ICD10 | I21.2 Acute transmural myocardial infarction of other sites |
| ICD10 | I21.3 Acute transmural myocardial infarction of unspecified site |
| ICD10 | I21.4 Acute subendocardial myocardial infarction |
| ICD10 | I21.9 Acute myocardial infarction, unspecified |
| ICD10 | I22 Subsequent myocardial infarction |
| ICD10 | I22.0 Subsequent myocardial infarction of anterior wall |
| ICD10 | I22.1 Subsequent myocardial infarction of inferior wall |
| ICD10 | I22.8 Subsequent myocardial infarction of other sites |
| ICD10 | I22.9 Subsequent myocardial infarction of unspecified site |
| ICD10 | I23 Certain current complications following acute myocardial infarction |
| ICD10 | I23.0 Haemopericardium as current complication following acute myocardial infarction |
| ICD10 | I23.1 Atrial septal defect as current complication following acute myocardial infarction |
| ICD10 | I23.2 Ventricular septal defect as current complication following acute myocardial infarction |
| ICD10 | I23.3 Rupture of cardiac wall without haemopericardium as current complication following acute myocardial infarction |
| ICD10 | I23.4 Rupture of chordae tendineae as current complication following acute myocardial infarction |
| ICD10 | I23.5 Rupture of papillary muscle as current complication following acute myocardial infarction |
| ICD10 | I23.6 Thrombosis of atrium, auricular appendage and ventricle as current complications following acute myocardial infarction |
| ICD10 | I23.8 Other current complications following acute myocardial infarction |

| | |
|---|---|
| ICD10 | I24 Other acute ischaemic heart diseases |
| ICD10 | I24.0 Coronary thrombosis not resulting in myocardial infarction |
| ICD10 | I24.1 Dressler's syndrome |
| ICD10 | I24.8 Other forms of acute ischaemic heart disease |
| ICD10 | I24.9 Acute ischaemic heart disease, unspecified |
| ICD10 | I25.1 Atherosclerotic heart disease |
| ICD10 | I25.2 Old myocardial infarction |
| ICD10 | I25.5 Ischaemic cardiomyopathy |
| ICD10 | I25.6 Silent myocardial ischaemia |
| ICD10 | I25.8 Other forms of chronic ischaemic heart disease |
| ICD10 | I25.9 Chronic ischaemic heart disease, unspecified |
| OPCS4 | K40 Saphenous vein graft replacement of coronary artery |
| OPCS4 | K40.1 Saphenous vein graft replacement of one coronary artery |
| OPCS4 | K40.2 Saphenous vein graft replacement of two coronary arteries |
| OPCS4 | K40.3 Saphenous vein graft replacement of three coronary arteries |
| OPCS4 | K40.4 Saphenous vein graft replacement of four or more coronary arteries |
| OPCS4 | K40.8 Other specified saphenous vein graft replacement of coronary artery |
| OPCS4 | K40.9 Unspecified saphenous vein graft replacement of coronary artery |
| OPCS4 | K41 Other autograft replacement of coronary artery |
| OPCS4 | K41.1 Autograft replacement of one coronary artery NEC |
| OPCS4 | K41.2 Autograft replacement of two coronary arteries NEC |
| OPCS4 | K41.3 Autograft replacement of three coronary arteries NEC |
| OPCS4 | K41.4 Autograft replacement of four or more coronary arteries NEC |
| OPCS4 | K41.8 Other specified other autograft replacement of coronary artery |
| OPCS4 | K41.9 Unspecified other autograft replacement of coronary artery |
| OPCS4 | K42 Allograft replacement of coronary artery |
| OPCS4 | K42.1 Allograft replacement of one coronary artery |
| OPCS4 | K42.2 Allograft replacement of two coronary arteries |
| OPCS4 | K42.3 Allograft replacement of three coronary arteries |
| OPCS4 | K42.4 Allograft replacement of four or more coronary arteries |
| OPCS4 | K42.8 Other specified allograft replacement of coronary artery |
| OPCS4 | K42.9 Unspecified allograft replacement of coronary artery |
| OPCS4 | K43 Prosthetic replacement of coronary artery |
| OPCS4 | K43.1 Prosthetic replacement of one coronary artery |
| OPCS4 | K43.2 Prosthetic replacement of two coronary arteries |
| OPCS4 | K43.3 Prosthetic replacement of three coronary arteries |
| OPCS4 | K43.4 Prosthetic replacement of four or more coronary arteries |
| OPCS4 | K43.8 Other specified prosthetic replacement of coronary artery |
| OPCS4 | K43.9 Unspecified prosthetic replacement of coronary artery |

| | |
|---|---|
| OPCS4 | K44 Other replacement of coronary artery |
| OPCS4 | K44.1 Replacement of coronary arteries using multiple methods |
| OPCS4 | K44.2 Revision of replacement of coronary artery |
| OPCS4 | K44.8 Other specified other replacement of coronary artery |
| OPCS4 | K44.9 Unspecified other replacement of coronary artery |
| OPCS4 | K45.1 Double anastomosis of mammary arteries to coronary arteries |
| OPCS4 | K45.2 Double anastomosis of thoracic arteries to coronary arteries NEC |
| OPCS4 | K45.3 Anastomosis of mammary artery to left anterior descending coronary artery |
| OPCS4 | K45.4 Anastomosis of mammary artery to coronary artery NEC |
| OPCS4 | K45.5 Anastomosis of thoracic artery to coronary artery NEC |
| OPCS4 | K45.6 Revision of connection of thoracic artery to coronary artery |
| OPCS4 | K45.8 Other specified connection of thoracic artery to coronary artery |
| OPCS4 | K45.9 Unspecified connection of thoracic artery to coronary artery |
| OPCS4 | K46 Other bypass of coronary artery |
| OPCS4 | K46.1 Double implantation of mammary arteries into heart |
| OPCS4 | K46.2 Double implantation of thoracic arteries into heart NEC |
| OPCS4 | K46.3 Implantation of mammary artery into heart NEC |
| OPCS4 | K46.4 Implantation of thoracic artery into heart NEC |
| OPCS4 | K46.5 Revision of implantation of thoracic artery into heart |
| OPCS4 | K46.8 Other specified other bypass of coronary artery |
| OPCS4 | K46.9 Unspecified other bypass of coronary artery |
| OPCS4 | K49.1 Percutaneous transluminal balloon angioplasty of one coronary artery |
| OPCS4 | K49.2 Percutaneous transluminal balloon angioplasty of multiple coronary arteries |
| OPCS4 | K49.3 Percutaneous transluminal balloon angioplasty of bypass graft of coronary artery |
| OPCS4 | K49.4 Percutaneous transluminal cutting balloon angioplasty of coronary artery |
| OPCS4 | K49.8 Other specified transluminal balloon angioplasty of coronary artery |
| OPCS4 | K49.9 Unspecified transluminal balloon angioplasty of coronary artery |
| OPCS4 | K50.1 Percutaneous transluminal laser coronary angioplasty |
| OPCS4 | K50.2 Percutaneous transluminal coronary thrombolysis using streptokinase |
| OPCS4 | K50.4 Percutaneous transluminal atherectomy of coronary artery |
| OPCS4 | K75.1 Percutaneous transluminal balloon angioplasty and insertion of 1-2 drug-eluting stents into coronary artery |
| OPCS4 | K75.2 Percutaneous transluminal balloon angioplasty and insertion of 3 or more drug-eluting stents into coronary artery |
| OPCS4 | K75.3 Percutaneous transluminal balloon angioplasty and insertion of 1-2 stents into coronary artery |
| OPCS4 | K75.4 Percutaneous transluminal balloon angioplasty and insertion of 3 or more stents into coronary artery NEC |

| | |
|---|---|
| OPCS4 | K75.8 Other specified percutaneous transluminal balloon angioplasty and insertion of stent into coronary artery |
| OPCS4 | K75.9 Unspecified percutaneous transluminal balloon angioplasty and insertion of stent into coronary artery |
| Self Report | heart attack/myocardial infarction |
| Operation code | coronary angioplasty (ptca) +/- stent |
| Operation code | coronary artery bypass grafts (cabg) |
| Operation code | triple heart bypass |
| Vascular/heart problems diagnosed by doctor | Heart attack |

Heart Failure

| **Source** | **Code** |
|---|---|
| ICD10 | I11.0 Hypertensive heart disease with (congestive) heart failure |
| ICD10 | I13.0 Hypertensive heart and renal disease with (congestive) heart failure |
| ICD10 | I13.2 Hypertensive heart and renal disease with both (congestive) heart failure and renal failure |
| ICD10 | I25.5 Ischaemic cardiomyopathy |
| ICD10 | I42.0 Dilated cardiomyopathy |
| ICD10 | I42.5 Other restrictive cardiomyopathy |
| ICD10 | I42.8 Other cardiomyopathies |
| ICD10 | I42.9 Cardiomyopathy, unspecified |
| ICD10 | I50 Heart failure |
| ICD10 | I50.0 Congestive heart failure |
| ICD10 | I50.1 Left ventricular failure |
| ICD10 | I50.9 Heart failure, unspecified |
| ICD9 | 4254 Other primary cardiomyopathies |
| ICD9 | 4280 Congestive heart failure |
| ICD9 | 4281 Left heart failure |
| Self Report | heart failure/pulmonary odema |
| Self Report | cardiomyopathy |
| ICD10 | I42.1 Obstructive hypertrophic cardiomyopathy |
| ICD10 | I42.2 Other hypertrophic cardiomyopathy |
| Self Report | hypertrophic cardiomyopathy (hcm / hocm) |

Aortic Stenosis

| **Source** | **Code** |
|---|---|

| | |
|---|---|
| ICD10 | I06.0 Rheumatic aortic stenosis |
| ICD10 | I06.2 Rheumatic aortic stenosis with insufficiency |
| ICD10 | I35.0 Aortic (valve) stenosis |
| ICD10 | I35.2 Aortic (valve) stenosis with insufficiency |
| Self Report | aortic stenosis |

Stroke

| Source | Code |
|---|---|
| Source of stroke report | Self-reported only |
| Source of stroke report | Hospital admission |
| Source of stroke report | Death only |

Hypertension

| Source | Code |
|---|---|
| ICD10 | I10 Essential (primary) hypertension |
| ICD10 | I11 Hypertensive heart disease |
| ICD10 | I11.0 Hypertensive heart disease with (congestive) heart failure |
| ICD10 | I11.9 Hypertensive heart disease without (congestive) heart failure |
| ICD10 | I12 Hypertensive renal disease |
| ICD10 | I12.0 Hypertensive renal disease with renal failure |
| ICD10 | I12.9 Hypertensive renal disease without renal failure |
| ICD10 | I13 Hypertensive heart and renal disease |
| ICD10 | I13.0 Hypertensive heart and renal disease with (congestive) heart failure |
| ICD10 | I13.1 Hypertensive heart and renal disease with renal failure |
| ICD10 | I13.2 Hypertensive heart and renal disease with both (congestive) heart failure and renal failure |
| ICD10 | I13.9 Hypertensive heart and renal disease, unspecified |
| ICD10 | I15 Secondary hypertension |
| ICD10 | I15.0 Renovascular hypertension |
| ICD10 | I15.1 Hypertension secondary to other renal disorders |
| ICD10 | I15.2 Hypertension secondary to endocrine disorders |
| ICD10 | I15.8 Other secondary hypertension |
| ICD10 | I15.9 Seconday hypertension, unspecified |
| ICD9 | 401 Essential hypertension |
| ICD9 | 4010 Essential hypertension, specified as malignant |
| ICD9 | 4011 Essential hypertension, specified as benign |
| ICD9 | 4019 Essential hypertension, not specified as malignant or benign |
| ICD9 | 402 Hypertensive heart disease |

| | |
|---|---|
| ICD9 | 4020 Hypertensive heart disease, specified as malignant |
| ICD9 | 4021 Hypertensive heart disease, specified as benign |
| ICD9 | 4029 Hypertensive heart disease, not specified as malignant or benign |
| ICD9 | 403 Hypertensive renal disease |
| ICD9 | 4030 Hypertensive renal disease, specified as malignant |
| ICD9 | 4031 Hypertensive renal disease, specified as benign |
| ICD9 | 4039 Hypertensive renal disease, not specified as malignant or benign |
| ICD9 | 404 Hypertensive heart and renal disease |
| ICD9 | 4040 Hypertensive heart and renal disease, specified as malignant |
| ICD9 | 4041 Hypertensive heart and renal disease, specified as benign |
| ICD9 | 4049 Hypertensive heart and renal disease, not specified as malignant or benign |
| ICD9 | 405 Secondary hypertension |
| ICD9 | 4050 Secondary hypertension, specified as malignant |
| ICD9 | 4051 Secondary hypertension, specified as benign |
| ICD9 | 4059 Secondary hypertension, not specified as malignant or benign |
| Self Report | hypertension |
| Self Report | essential hypertension |
| Vascular/heart problems diagnosed by doctor | High blood pressure |

Atrial Fibrillation

| **Type** | **Code** |
|---|---|
| ICD10 | I48 Atrial fibrillation and flutter |
| ICD10 | I48.0 Paroxysmal atrial fibrillation |
| ICD10 | I48.1 Persistent atrial fibrillation |
| ICD10 | I48.2 Chronic atrial fibrillation |
| ICD10 | I48.3 Typical atrial flutter |
| ICD10 | I48.4 Atypical atrial flutter |
| OPCS4 | K62.1 Percutaneous transluminal ablation of pulmonary vein to left atrium conducting system |
| OPCS4 | K62.2 Percutaneous transluminal ablation of atrial wall for atrial flutter |
| OPCS4 | K62.3 Percutaneous transluminal ablation of conducting system of heart for atrial flutter NEC |
| Self Report | atrial fibrillation |
| Self Report | atrial flutter |

Diabetes Mellitus

| Source | Code |
|---|---|
| ICD10 | E10 Insulin-dependent diabetes mellitus |
| ICD10 | E10.0 With coma |
| ICD10 | E10.1 With ketoacidosis |
| ICD10 | E10.2 With renal complications |
| ICD10 | E10.3 With ophthalmic complications |
| ICD10 | E10.4 With neurological complications |
| ICD10 | E10.5 With peripheral circulatory complications |
| ICD10 | E10.6 With other specified complications |
| ICD10 | E10.7 With multiple complications |
| ICD10 | E10.8 With unspecified complications |
| ICD10 | E10.9 Without complications |
| Self Report | type 1 diabetes |
| ICD10 | E11 Non-insulin-dependent diabetes mellitus |
| ICD10 | E11.0 With coma |
| ICD10 | E11.1 With ketoacidosis |
| ICD10 | E11.2 With renal complications |
| ICD10 | E11.3 With ophthalmic complications |
| ICD10 | E11.4 With neurological complications |
| ICD10 | E11.5 With peripheral circulatory complications |
| ICD10 | E11.6 With other specified complications |
| ICD10 | E11.7 With multiple complications |
| ICD10 | E11.8 With unspecified complications |
| ICD10 | E11.9 Without complications |
| Self Report | type 2 diabetes |

Hypercholesterolemia

| **Source** | **Code** |
|---|---|
| ICD10 | E10 Insulin-dependent diabetes mellitus |
| ICD10 | E10.0 With coma |
| ICD10 | E10.1 With ketoacidosis |
| ICD10 | E10.2 With renal complications |
| ICD10 | E10.3 With ophthalmic complications |
| ICD10 | E10.4 With neurological complications |
| ICD10 | E10.5 With peripheral circulatory complications |
| ICD10 | E10.6 With other specified complications |
| ICD10 | E10.7 With multiple complications |
| ICD10 | E10.8 With unspecified complications |

| | |
|---|---|
| ICD10 | E10.9 Without complications |
| Self Report | type 1 diabetes |
| ICD10 | E11 Non-insulin-dependent diabetes mellitus |
| ICD10 | E11.0 With coma |
| ICD10 | E11.1 With ketoacidosis |
| ICD10 | E11.2 With renal complications |
| ICD10 | E11.3 With ophthalmic complications |
| ICD10 | E11.4 With neurological complications |
| ICD10 | E11.5 With peripheral circulatory complications |
| ICD10 | E11.6 With other specified complications |
| ICD10 | E11.7 With multiple complications |
| ICD10 | E11.8 With unspecified complications |
| ICD10 | E11.9 Without complications |
| Self Report | type 2 diabetes |

Cancer

| **Source** | **Code** |
|---|---|
| Cancer record origin | Originating from England/Wales |
| Cancer record origin | Hospital Episode Statistics from England |
| Cancer record origin | National Cancer Intelligence Network |
| Cancer record origin | Patient Episode Database for Wales |
| Cancer record origin | Originating from Scotland |
| Cancer record origin | Scottish Morbidity Records |

Abbreviations
ICD, International Classification of Diseases; OPCS4, OPCS Classifications of Interventions and Procedures, version 4

**Supplemental Table 2: Comparative Assessment of 20 Polygenic Risk Scores for Notch Smoothness**

| Linear Regression Model | PRS tuning parameters | | Linear Regression Model Fit in Validation Cohort | |
|---|---|---|---|---|
| | LD ($r^2$) | P-value | $R^2$ | Improvement in $R^2$ over clinical model |
| Clinical Model: notch smoothness ~ age + sex + genotyping array + PCs 1-12 | - | - | 0.0808 | - |
| Clinical model + PRS1 | 0.8 | $5x10^{-8}$ | 0.0828 | 0.002 |
| Clinical model + PRS2 | 0.6 | $5x10^{-8}$ | 0.0828 | 0.002 |
| Clinical model + PRS3 | 0.4 | $5x10^{-8}$ | 0.0827 | 0.002 |
| Clinical model + PRS4 | 0.2 | $5x10^{-8}$ | 0.0827 | 0.002 |
| Clinical model + PRS5 | 0.05 | $5x10^{-8}$ | 0.0827 | 0.002 |
| Clinical model + PRS6 | 0.8 | $5x10^{-6}$ | 0.0828 | 0.002 |
| Clinical model + PRS7 | 0.6 | $5x10^{-6}$ | 0.0828 | 0.002 |
| Clinical model + PRS8 | 0.4 | $5x10^{-6}$ | 0.0826 | 0.002 |
| Clinical model + PRS9 | 0.2 | $5x10^{-6}$ | 0.0827 | 0.002 |
| Clinical model + PRS10 | 0.05 | $5x10^{-6}$ | 0.0827 | 0.002 |
| Clinical model + PRS11 | 0.8 | $5x10^{-4}$ | 0.0832 | 0.002 |
| Clinical model + PRS12 | 0.6 | $5x10^{-4}$ | 0.0830 | 0.002 |
| Clinical model + PRS13 | 0.4 | $5x10^{-4}$ | 0.0825 | 0.002 |
| Clinical model + PRS14 | 0.2 | $5x10^{-4}$ | 0.0822 | 0.001 |
| Clinical model + PRS15 | 0.05 | $5x10^{-4}$ | 0.0821 | 0.001 |
| Clinical model + PRS16 | 0.8 | 0.05 | 0.0836 | 0.003 |
| **Clinical model + PRS17** | **0.6** | **0.05** | **0.0836** | **0.003** |
| Clinical model + PRS18 | 0.4 | 0.05 | 0.0833 | 0.003 |
| Clinical model + PRS19 | 0.2 | 0.05 | 0.0828 | 0.002 |
| Clinical model + PRS20 | 0.05 | 0.05 | 0.0825 | 0.002 |

Bolding indicates top performing score in the validation cohort.

**Supplemental Table 3: Baseline Characteristics of Participants With and Without Binary Absent Notch**

| | No Absent Notch | Absent Notch | |
|---|---|---|---|
| | n=144501 | n=25286 | p-value |
| Age | 56.53 (8.20) | 61.51 (6.36) | <0.001 |
| Men | 69124 (47.8) | 8637 (34.2) | <0.001 |
| Heart rate | 68.57 (11.26) | 70.09 (13.12) | <0.001 |
| BMI | 27.35 (4.79) | 28.11 (4.99) | <0.001 |
| SBP | 139.26 (19.40) | 147.36 (20.56) | <0.001 |
| DBP | 82.16 (10.66) | 82.74 (10.79) | <0.001 |
| Prevalent Conditions | | | |
| Ever Smoking | 84939 (59.1) | 15741 (62.8) | <0.001 |
| Hypercholesterolemia | 21642 (15.0) | 5800 (22.9) | <0.001 |
| Diabetes mellitus | 4058 (2.8) | 1135 (4.5) | <0.001 |
| Heart Failure | 798 (0.6) | 207 (0.8) | <0.001 |
| MI/CAD | 5597 (3.9) | 1608 (6.4) | <0.001 |
| Aortic stenosis | 204 (0.1) | 76 (0.3) | <0.001 |
| Stroke | 1934 (1.3) | 612 (2.4) | <0.001 |
| Atrial fibrillation | 2270 (1.6) | 685 (2.7) | <0.001 |
| Any Cancer | 12477 (8.6) | 2757 (10.9) | <0.001 |
| PPG Features | | | |
| Position of Shoulder | 20.28 (5.46) | 26.08 (3.49) | <0.001 |

| | | | |
|---|---|---|---|
| Position of Peak | 21.56 (5.38) | 26.39 (3.39) | <0.001 |
| Position of Notch | 43.48 (6.25) | 43.97 (6.90) | <0.001 |
| Wave Reflection Index | 66.72 (31.42) | 73.46 (35.37) | <0.001 |
| Arterial Stiffness Index | 8.95 (3.59) | 11.54 (5.51) | <0.001 |

**Supplemental Table 4: Multivariable Adjusted Association of Binary Absent Notch & Notch Smoothness with Prevalent Cardiovascular Disease at Baseline**

| | Binary Absent Notch | | Notch Smoothness | |
|---|---|---|---|---|
| | Adj OR (95% CI) | P-Value | Adj OR per SD (95% CI) | P-Value |
| Hypertension | 1.19 (1.15-1.23) | 2.6E-24 | 1.07 (1.06-1.08 | 1.7E-27 |
| MI/CAD | 1.23 (1.15-1.32) | 7.3E-10 | 1.10 (1.07-1.13) | 4.8E-12 |
| Heart Failure | 1.18 (1.00-1.38) | 0.05 | 1.08 (1.01-1.15) | 0.03 |
| Stroke | 1.43 (1.29-1.58) | 1.1E-12 | 1.18 (1.13-1.23) | 2.0E-15 |
| Aortic Stenosis | 1.3 (0.98-1.73) | 0.07 | 1.16 (1.03-1.30) | 0.01 |
| Atrial Fibrillation | 1.42 (1.29-1.56) | 9.1E-14 | 1.17 (1.13-1.21) | 6.7E-16 |
| Any Cancer | 0.94 (0.89-0.98) | 0.01 | 0.97 (0.95-0.98) | 1.7E-04 |

Covariates included: age, sex, HR, BMI, SBP, DBP, prevalent type II DM, hypercholesterolemia, and ever smoking

**Supplemental Table 5: Variants Associated with Binary Absent Notch in Multi-Ancestry Population**

| Rsid | Chr | Position (hg18) | Nearest Gene | Effect Allele | Allele Frequency (%) | OR (95% CI) | P Value |
|---|---|---|---|---|---|---|---|
| 2:145690116_CT_C | 2 | 145690116 | *TEX41* | C | 36% | 1.07 (1.04,1.09) | 2.24E-08 |
| rs7652774 | 3 | 141579311 | *ATP1B3* | C | 38% | 1.07 (1.05,1.1) | 3.81E-10 |
| rs11977526 | 7 | 46008110 | *IGFBP3* | A | 41% | 0.91 (0.89,0.93) | 1.02E-17 |
| rs1926034 | 10 | 104829102 | *CNNM2 / NT5C2* | A | 38% | 0.91 (0.89,0.93) | 5.53E-18 |
| rs11570072 | 11 | 47365276 | *MYBPC3* | G | 17% | 0.91 (0.88,0.93) | 4.89E-11 |
| rs227424 | 14 | 70456804 | *SMOC1* | A | 43% | 0.93 (0.91,0.95) | 3.14E-10 |

Odds ratio indicates expected change in odds of binary absent notch per allele.

Abbreviations: Chr, chromosome; CI, confidence interval; OR, odds ratio.

**Supplemental Table 6: Variants Associated with Notch Smoothness in Participants of European Ancestry**

| Rsid | Chr | Position (hg18) | Nearest Gene | Effect Allele | Allele Frequency (%) | Beta (95% CI) | P Value |
|---|---|---|---|---|---|---|---|
| rs13386034 | 2 | 145759899 | *TEX41* | C | 25% | 0.025 (0.016,0.033) | 4.12E-08 |
| rs7652774 | 3 | 141579311 | *ATP1B3* | C | 39% | 0.024 (0.016,0.032) | 1.40E-09 |
| rs744892 | 3 | 14871766 | *FGD5* | A | 12% | 0.035 (0.024,0.047) | 3.20E-09 |
| rs6890990 | 5 | 122599707 | *PRDM6 / CEP120* | G | 20% | -0.027 (-0.037,-0.017) | 4.28E-08 |
| rs9349379 | 6 | 12903957 | *PHACTR1* | G | 40% | -0.026 (-0.034,-0.018) | 4.53E-11 |
| rs11977526 | 7 | 46008110 | *IGFBP3* | A | 40% | -0.03 (-0.038,-0.023) | 1.67E-14 |
| rs1926034 | 10 | 104829102 | *CNNM2 / NT5C2* | A | 37% | -0.038 (-0.046,-0.03) | 1.13E-21 |
| rs4751648 | 10 | 116132246 | *AFAP1L2* | C | 45% | 0.025 (0.017,0.032) | 1.91E-10 |
| rs2269434 | 11 | 47360412 | *MYBPC3* | C | 35% | -0.024 (-0.032,-0.016) | 1.88E-09 |
| rs10770612 | 12 | 20230639 | *LOC100506393* | G | 20% | -0.028 (-0.038,-0.019) | 4.78E-09 |
| rs4773173 | 13 | 111025118 | *COL4A2* | G | 34% | -0.023 (-0.031,-0.015) | 2.19E-08 |
| rs227439 | 14 | 70451780 | *SMOC1* | G | 42% | -0.024 (-0.032,-0.016) | 1.56E-09 |
| rs17514846 | 15 | 91416550 | *FURIN* | A | 47% | 0.023 (0.016,0.031) | 2.15E-09 |

Beta coefficient indicates expected increase in standardized continuous trait per allele. European ancestry participants (n=123,644) ) were defined as self-identifying White participants after exclusion of outliers in 3 pairs of principal components (1-2, 3-4, and 5-6) using the R package aberrant (lambda=40).
Abbreviations: Chr, chromosome; CI, confidence interval

**Supplemental Table 7: Association of Notch Smoothness with Incident Cardiovascular Events in Participants Free of Cardiovascular Disease at Baseline**

| | | Adjusted for Age & Sex | | Adjusted for Clinical Covariates | | Adjusted for Clinical Covariates & Arterial Stiffness Index | |
|---|---|---|---|---|---|---|---|
| Outcome | Events | HR per SD (95% CI) | P-Value | HR per SD (95% CI) | P-Value | HR per SD (95% CI) | P-Value |
| Hypertension | 12084 | 1.15 (1.13-1.17) | 7.3E-51 | 1.06 (1.04-1.08) | 7.0E-11 | 1.03 (1.01-1.05) | 0.01 |
| MI/CAD | 8546 | 1.14 (1.11-1.16) | 1.1E-31 | 1.08 (1.06-1.11) | 8.6E-13 | 1.06 (1.04-1.09) | 7.2E-07 |
| Heart Failure | 2560 | 1.17 (1.12-1.21) | 1.4E-15 | 1.08 (1.04-1.13) | 4.0E-05 | 1.08 (1.04-1.13) | 2.9E-04 |
| Stroke | 1202 | 1.12 (1.06-1.18) | 1.1E-04 | 1.08 (1.02-1.15) | 0.01 | 1.08 (1.02-1.15) | 0.01 |
| Aortic Stenosis | 911 | 1.24 (1.16-1.31) | 3.5E-11 | 1.14 (1.07-1.21) | 8.8E-05 | 1.14 (1.07-1.22) | 1.5E-04 |
| Atrial Fibrillation | 6371 | 1.06 (1.03-1.08) | 1.7E-05 | 1.02 (0.99-1.04) | 0.13 | 1.03 (1-1.05) | 0.06 |
| Death | 6816 | 1.13 (1.11-1.16) | 5.2E-25 | 1.09 (1.07-1.12) | 5.0E-13 | 1.09 (1.06-1.11) | 6.9E-10 |
| Cancer | 11120 | 1 (0.98-1.02) | 0.90 | 1 (0.98-1.02) | 0.74 | 0.99 (0.96-1.01) | 0.16 |

Analysis of incident events was restricted to participants free of cardiovascular disease (MI/CAD, heart failure, stroke, aortic stenosis, or atrial fibrillation) at baseline (n-155,717). Participants with prevalent CVD (myocardial infarction or coronary artery disease, heart failure, stroke, aortic stenosis, or atrial fibrillation) or the outcome of interest at baseline were excluded. Clinical covariates: age, sex, heart rate, body mass index, systolic and diastolic blood pressure prevalent diabetes mellitus, hypercholesterolemia, and ever smoking. All results exclude patients with any of these conditions at baseline. CI, confidence interval; HR, hazard ratio; MI/CAD, myocardial infarction or coronary artery disease; SD, standard deviation.

**Supplemental Table 8: Association of Binary Absent Notch with Incident Cardiovascular Events in Participants Free of Cardiovascular Disease at Baseline**

| | | Adjusted Age/Sex | | Adjusted Clinical Covariates | | Adjusted for Clinical Covariates & Arterial Stiffness Index | |
|---|---|---|---|---|---|---|---|
| Outcome | Events | HR (95% CI) | P-Value | HR (95% CI) | P-Value | HR (95% CI) | P-Value |
| Hypertension | 12084 | 1.39 (1.33-1.46) | 3.8E-45 | 1.17 (1.11-1.22) | 1.9E-10 | 1.09 (1.04-1.15) | 6.9E-04 |
| MI/CAD | 8546 | 1.31 (1.24-1.38) | 9.5E-23 | 1.19 (1.13-1.26) | 7.2E-10 | 1.14 (1.08-1.21) | 6.7E-06 |
| Heart Failure | 2560 | 1.40 (1.27-1.53) | 1.7E-12 | 1.21 (1.10-1.33) | 7.8E-05 | 1.20 (1.08-1.32) | 4.5E-04 |
| Stroke | 1202 | 1.28 (1.12-1.48) | 4.0E-04 | 1.2 (1.04-1.38) | 0.01 | 1.19 (1.03-1.38) | 0.02 |
| Aortic Stenosis | 911 | 1.49 (1.28-1.74) | 2.4E-07 | 1.25 (1.07-1.46) | 4.1E-03 | 1.24 (1.06-1.46) | 8.0E-03 |
| Atrial Fibrillation | 6371 | 1.13 (1.06-1.2) | 1.2E-04 | 1.06 (0.99-1.13) | 0.08 | 1.07 (1-1.14) | 0.05 |
| Death | 6816 | 1.32 (1.25-1.4) | 5.8E-21 | 1.24 (1.17-1.32) | 1.2E-12 | 1.22 (1.14-1.3) | 5.6E-10 |
| Cancer | 11120 | 1.00 (0.95-1.05) | 0.88 | 0.99 (0.94-1.04) | 0.76 | 0.97 (0.92-1.03) | 0.32 |

Analysis of incident events was restricted to participants free of cardiovascular disease (MI/CAD, heart failure, stroke, aortic stenosis, or atrial fibrillation) at baseline (n=155,717). Participants with prevalent hypertension were also excluded for hypertension outcome only. Clinical covariates: age, sex, heart rate, body mass index, systolic and diastolic blood pressure prevalent diabetes mellitus, hypercholesterolemia, and ever smoking. All results exclude patients with any of these conditions at baseline. MI/CAD, myocardial infarction or coronary artery disease.

**Supplemental Table 9: C-Statistics for Incident Cardiovascular Disease With Binary Absent Notch and Notch Smoothness Traits**

| | Univariable | | Multivariable (with clinical covariates) | | |
|---|---|---|---|---|---|
| Univariate | Binary Absent Notch | Notch Smoothness | Clinical Only | Binary Absent Notch | Notch Smoothness |
| Hypertension | 0.538 | 0.566 | 0.757 | 0.758 | 0.758 |
| MI/CAD | 0.531 | 0.565 | 0.716 | 0.717 | 0.717 |
| Heart Failure | 0.546 | 0.581 | 0.749 | 0.749 | 0.749 |
| Stroke | 0.539 | 0.566 | 0.708 | 0.709 | 0.709 |
| Aortic Stenosis | 0.556 | 0.607 | 0.792 | 0.794 | 0.794 |
| Atrial Fibrillation | 0.531 | 0.559 | 0.745 | 0.745 | 0.745 |
| Cancer | 0.519 | 0.537 | 0.656 | 0.656 | 0.656 |
| Death | 0.540 | 0.569 | 0.717 | 0.718 | 0.718 |

C-statistics refer to Cox proportional hazards regression models restricted to patients free of cardiovascular disease (MI/CAD, heart failure, stroke, aortic stenosis, or atrial fibrillation) and the outcome of interest at baseline. Multivariable model includes the follow covariates: age, sex, body mass index, heart rate, systolic and diastolic blood pressure, ever smoking, prevalent hypercholesterolemia, and prevalent diabetes mellitus.

**Supplemental Table 10: Cross-Sectional Association of Notch Smoothness & Cardiac MRI Traits at Imaging Visit**

Notch Smoothness

| | Difference per SD Notch Smoothness (95% CI) | | |
|---|---|---|---|
| | Unadjusted | Adj. Age & Sex | Adj Age, Sex, S/DBP, HR, & BMI |
| LV End Diastolic Volume Indexed to BSA (ml/m$^2$) | -1.81 (-1.67,-1.95), p=6x10$^{-143}$ | -0.92 (-0.78,-1.05), p-2x10$^{-38}$ | -0.48 (-0.35,-0.61), p=7x10$^{-13}$ |
| LV End Systolic Volume Indexed to BSA (ml/m$^2$) | -1.05 (-0.97,-1.14), p=7x10$^{-133}$ | -0.57 (-0.49,-0.66), p=7x10$^{-43}$ | -0.39 (-0.31,-0.48), p=7x10$^{-20}$ |
| LV Ejection Fraction (%) | 0.59 (0.50,0.66), p=2x10$^{-70}$ | 0.36 (0.30,0.43), p=7x10$^{-29}$ | 0.33 (0.30,0.40), p<=3x10$^{-21}$ |

BMI, body mass index; BSA, body surface area; HR, heart rate; LV, left ventricle; S/DBP, systolic and diastolic blood pressure

Binary Absent Notch

| | Difference in Absent Notch (95% CI) | | |
|---|---|---|---|
| | Unadjusted | Adj. Age & Sex | Adj Age, Sex, S/DBP, HR, & BMI |
| LV End Diastolic Volume Indexed to BSA(ml/m$^2$) | -3.5 (-3.8,-3.1), p=6x10$^{-100}$ | -1.5 (-1.8,-1.2), p=1x10$^{-20}$ | -0.9 (-1.2,-0.6), p=6x10$^{-9}$ |
| LV End Systolic Volume Indexed to BSA (ml/m$^2$) | -2.1 (-2.3,-1.9), p=8x10$^{-100}$ | -1.0 (-1.2,-0.8), p=6x10$^{-26}$ | -0.7 (-0.9,-0.5), p=2x10$^{-13}$ |

| | | | |
|---|---|---|---|
| LV Ejection Fraction (%) | 1.2 (1.1,1.4), $p=6x10^{-60}$ | 0.7 (0.6,0.8), $p=4x10^{-21}$ | 0.6 (0.5,0.8), $p=5x10^{-15}$ |

BMI, body mass index; BSA, body surface area; HR, heart rate; LV, left ventricle; S/DBP, systolic and diastolic blood pressure

### Supplemental Table 11: Annotation of Genome Wide Significant Loci for Notch Smoothness

| Top SNP at Locus | Chr:Pos (hg18) | eQTL in Aorta or Left Ventricle | Nearest Gene | Closest Gene in TWAS (Aorta or LV) | Human disease in OMIM | Cardiovascular Effect of Gene Knockout |
|---|---|---|---|---|---|---|
| rs763944709 | chr2:145700572 | *ZEB2* (aorta) | *TEX41* | *ZEB2* | ZEB2 loss of function mutations cause Mowat-Wilson syndrome (microcephaly, mental retardation, hypertelorism, submucous cleft palate, and short stature (PMID: 11279515) | - |
| rs7652774 | chr3:141579311 | | *ATP1B3* | *ATP1B3* | - | - |
| rs744892 | chr3:14871766 | - | *FGD5* | *MRPS25* | MRPS25 mutations cause familial oxidative phosphorylation deficiency-50 causing mitochrondrial encephalopathy, consistent with role of the gene product (mitochrondrial ribosomal protein S25) | - |
| rs6890990 | chr5:122599707 | *PRDM6* (aorta) | *PRDM6 & CEP120* | - | PRDM6 missense mutations cause autosomal dominant nonsyndromic patent ductus arteriosus (PMID 27181681) | - |
| rs9349379 | chr6:12903957 | *PHACTR1* (aorta) | *PHACTR1* | *PHACTR1* | PHACTR1 heterozygous transversion mutations cause developmental and epileptic encephalopathy (PMID: 30256902 & 23033978) | CRISPR-edited stem cell-derived endothelial cells demonstrate that PHACTR1 locus (6p24) regulates expression of endothelin 1. a vasoconstrictor. |
| rs11977526 | chr7:46008110 | *IGFBP3* (left ventricle) | *IGBP3* | *IGBP3* | - | IGBP3-deficient mice demonstrate dose-dependent increase in oxygen-induced retinal vessel loss and decrease in vessel regrowth (PMID 17567756). |
| rs1537370 | chr9:22084310 | | *CDKN2B-AS1* | | | |
| rs1926034 | chr10:104829102 | *NT5C2, BORCS7 & AS3MT* (aorta) | *CNNM2 & NT5C2* | *NT5C2, BORCS7 & AS3MT* | NT5C2: homozygous nonsense mutations asociated with spastic paraplegia (PMID 24482476). | NT5C2 knockout mice demonstrate reduced body weight, adipose tissue, and insulin resistance (PMID: 30803894). |
| rs4751648 | chr10:116132246 | - | *AFAP1L2* | - | - | - |

| | | | | | | |
|---|---|---|---|---|---|---|
| rs2269434 | chr11:47360412 | *C1QTNF4* (aorta) | *MYBPC3* | - | Truncating mutations in MYBPC3 resulting in haploinsufficiency cause familial hypertrophic cardiomyopathy by augmentation of myosin contractility (PMID: 30674652) | - |
| rs28691743 | chr11:69797119 | | *ANO1* | | | |
| rs10770612 | chr12:20230639 | - | *LOC100506393* | - | - | - |
| rs9521719 | chr13:111017784 | - | *COL4A2* | - | Missense mutations in COL4A2 are associated with porencephaly, a neurological disorder characterized by fluid-filled cysts due to small vessel hemorrhage (PMID 22209246). | Mice with heterozygous G646D mutation in COL4A2 developed multifocal intracerebral hemorrhages (PMID: 24203695) and myopathy (PMID: 22209247). |
| rs227424 | chr14:70456804 | - | *SMOC1* | *SMOC1* | Homozygous mutations in SMOC1 cause micropthamia and limb abnormalities | - |
| rs6224 | chr15:91423543 | *FES* & *FURIN* (aorta) | *FURIN* | *FES* | - | - |

### Supplemental Figure 1: CONSORT Diagram

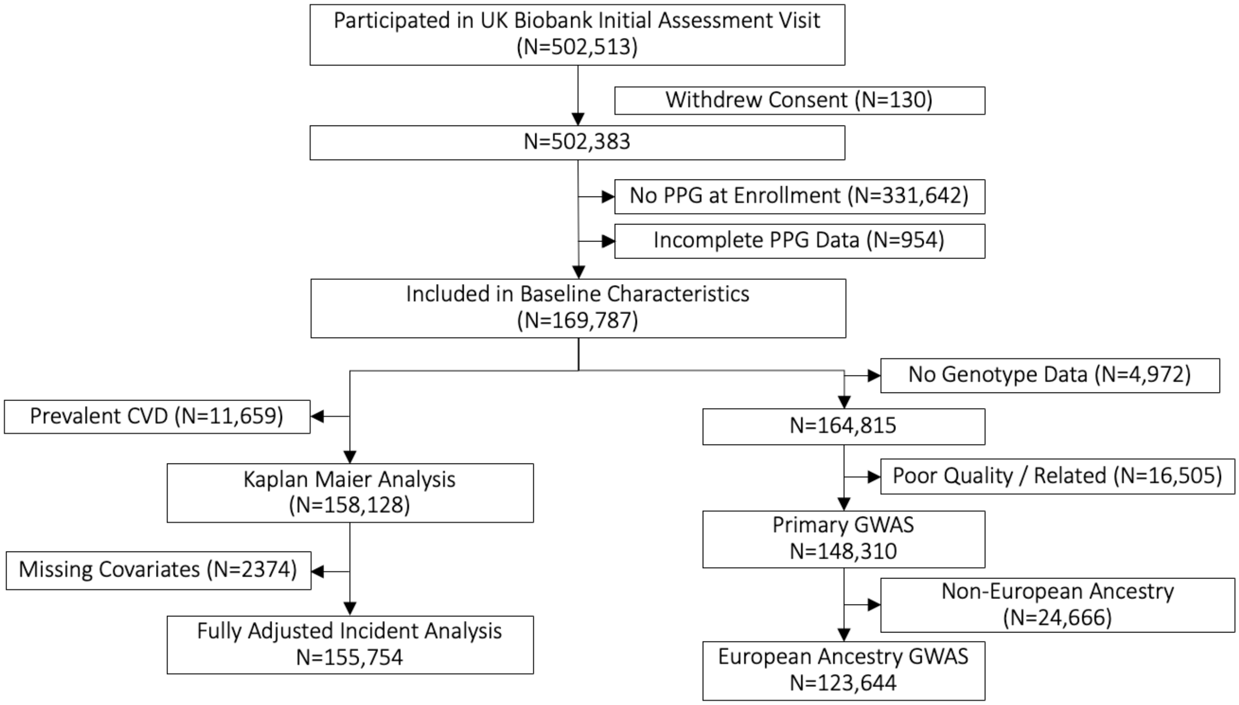

**Supplemental Figure 2: Q-Q Plot for Notch Smoothness GWAS**

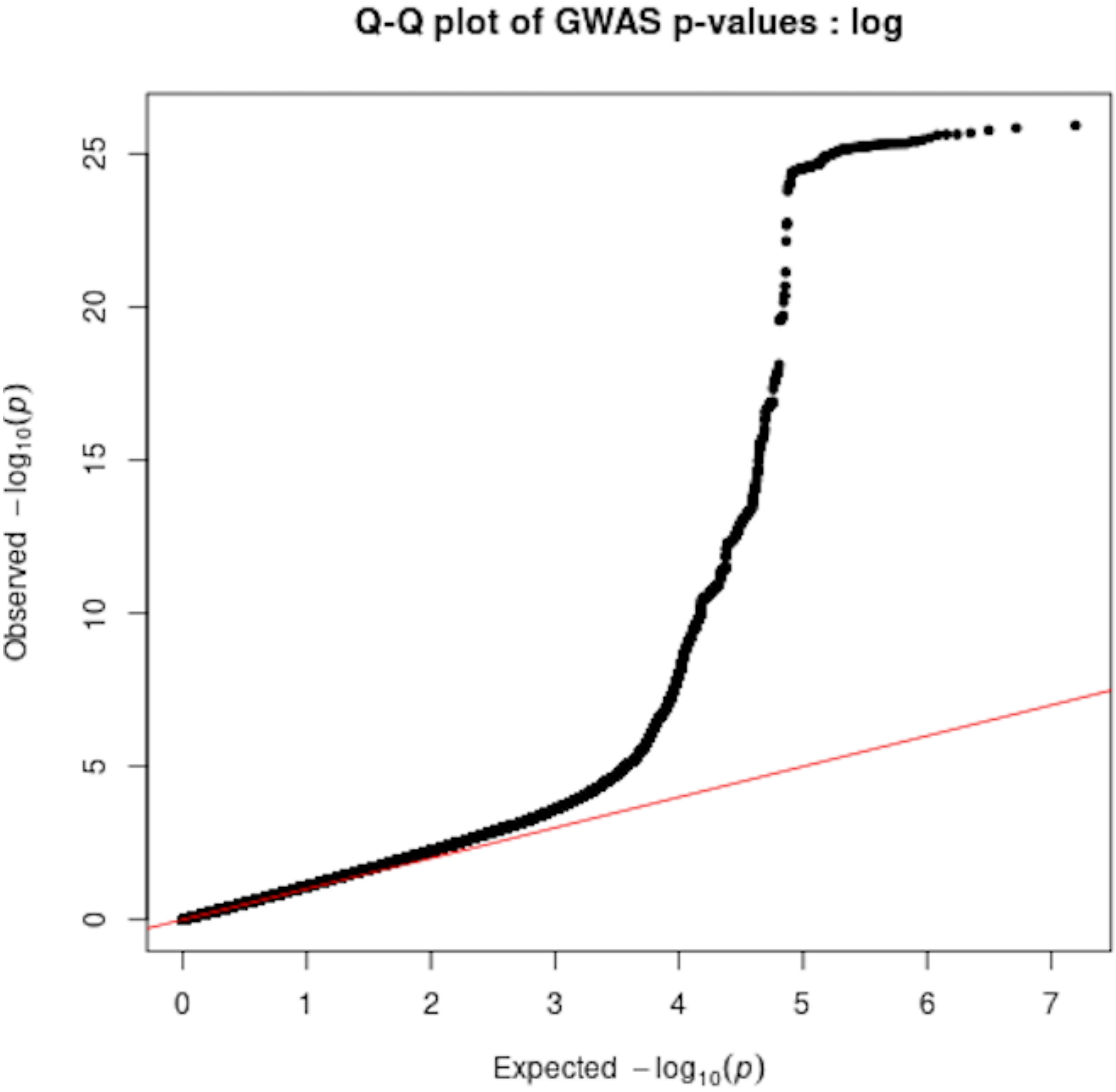

**Supplemental Figure 3: Locus Zoom Plots for Loci Associated with Notch Smoothness**

Loci with lead variant p-value < $5x10^{-8}$ are shown.

<u>Chromosome 2 (near *TEX41*)</u>

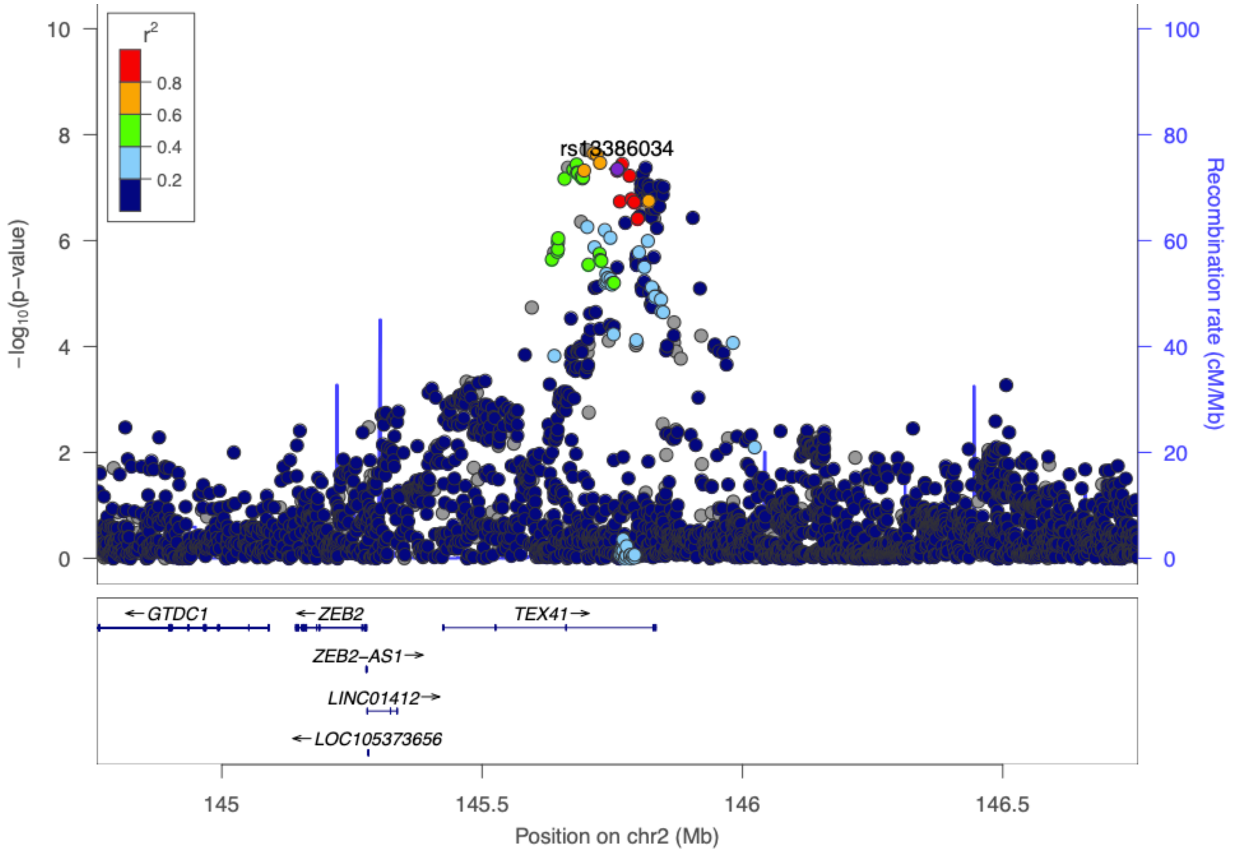

Chromosome 3 (near *ATP1B3*)

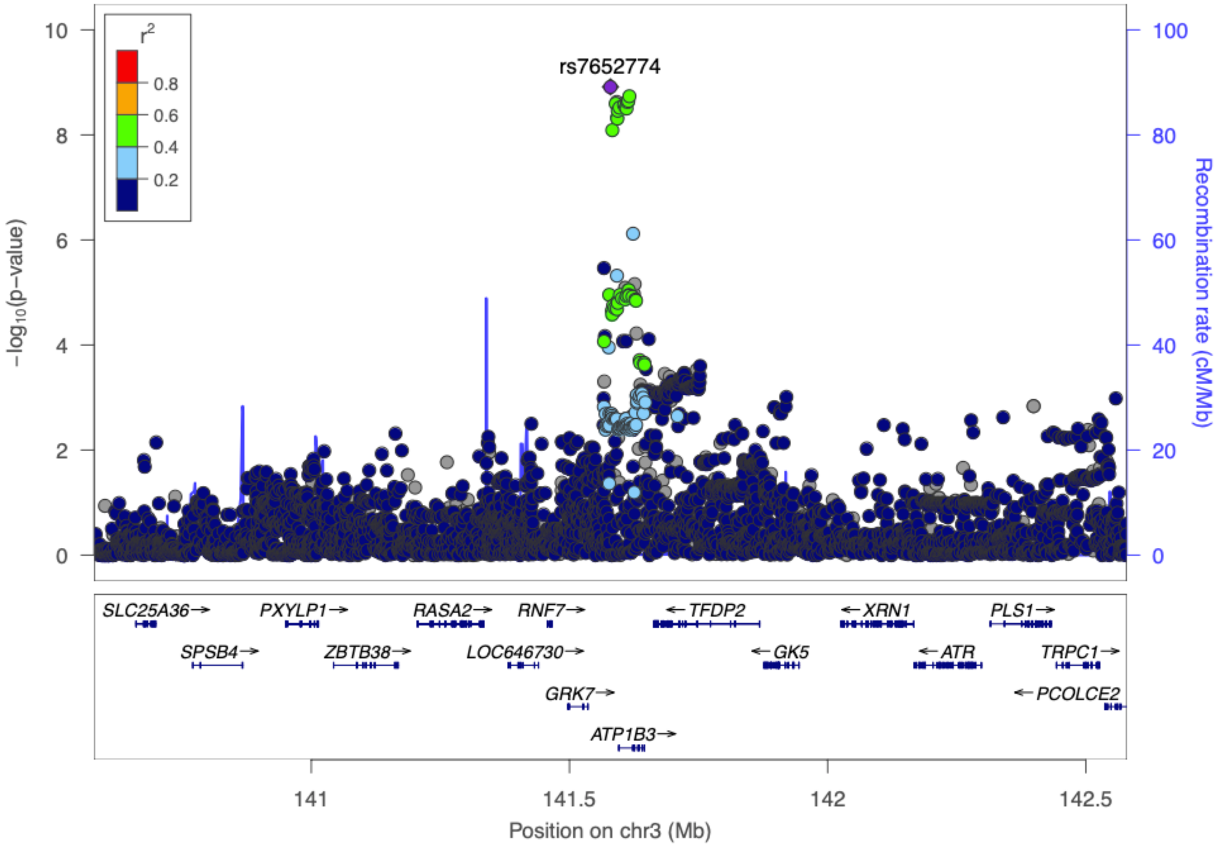

Chromosome 3 (near *FGD5*)

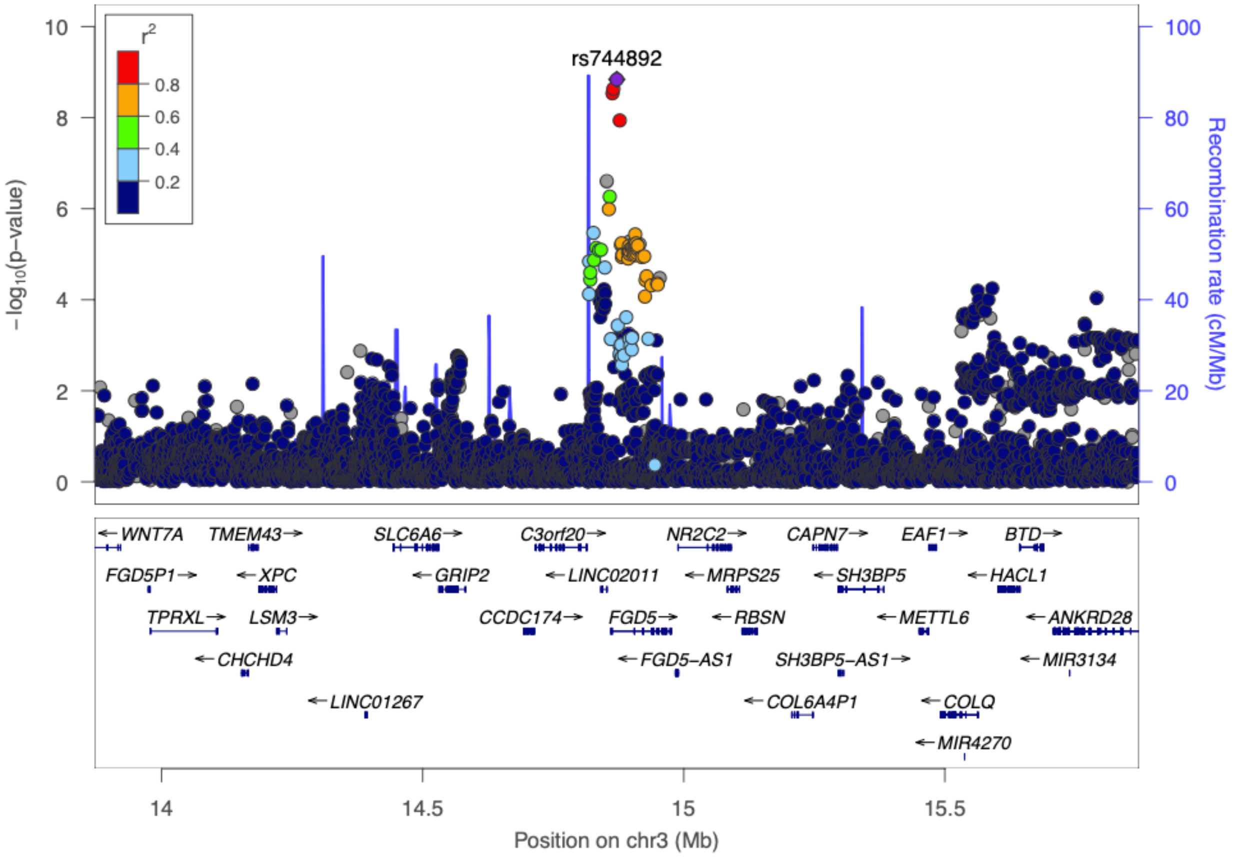

Chromosome 5 (near *PRDM6* & *CEP120*)

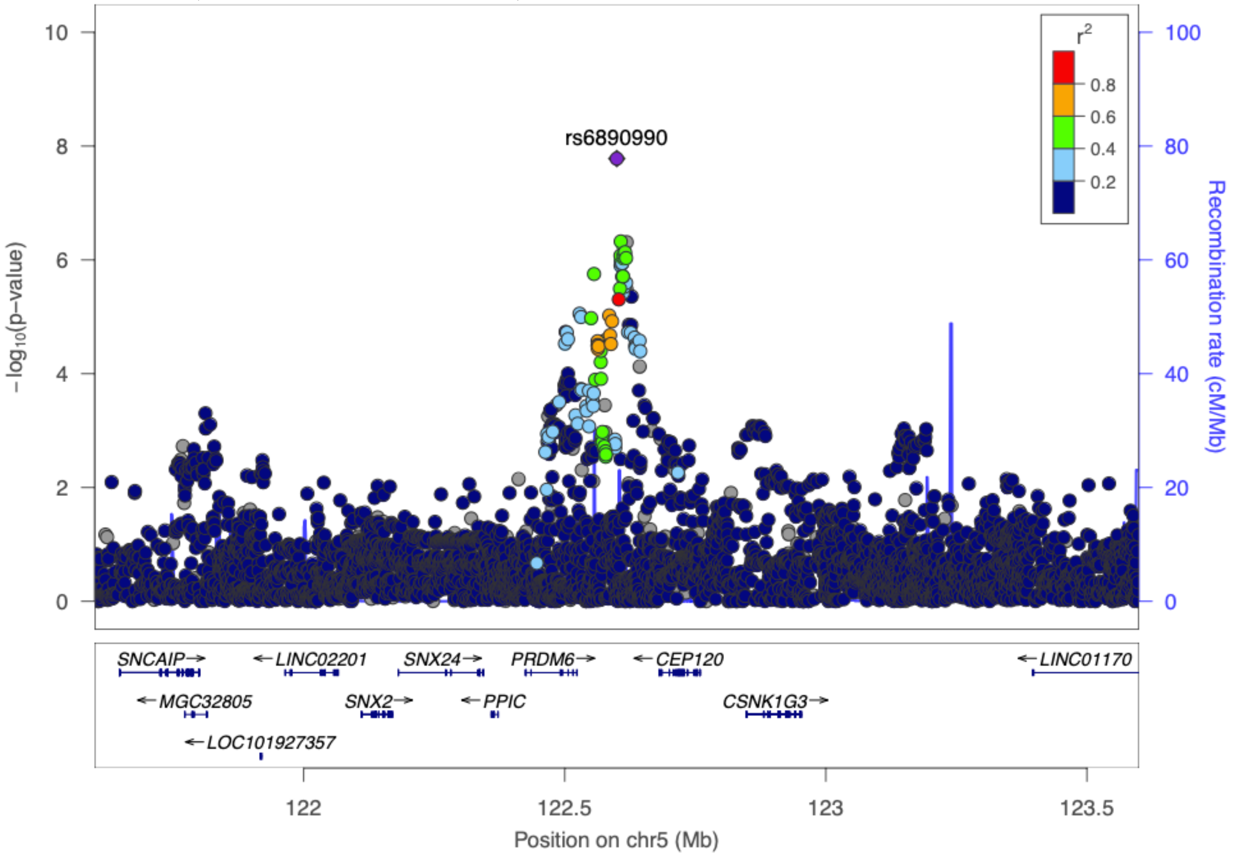

Chromosome 6 (near *PHACTR1*)

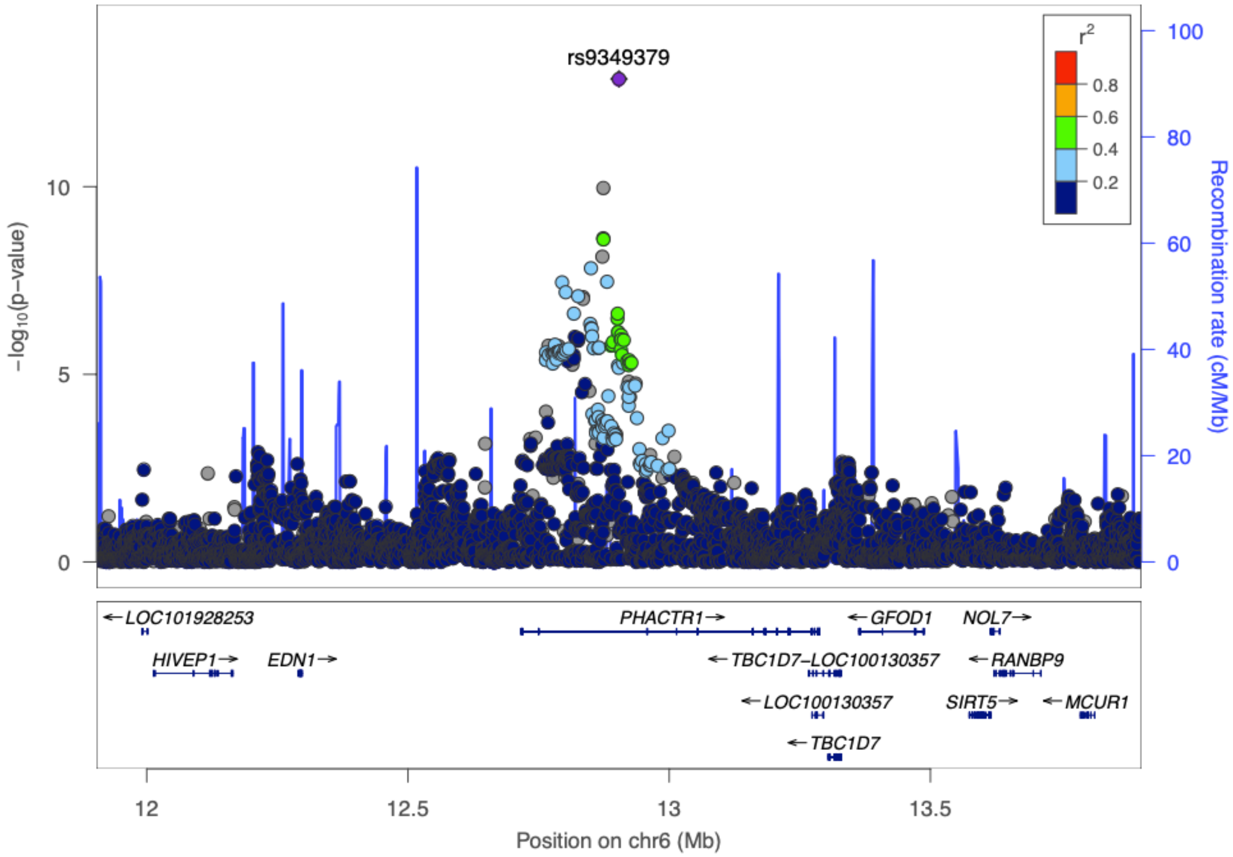

Chromosome 7 (near *IGFBP3*)

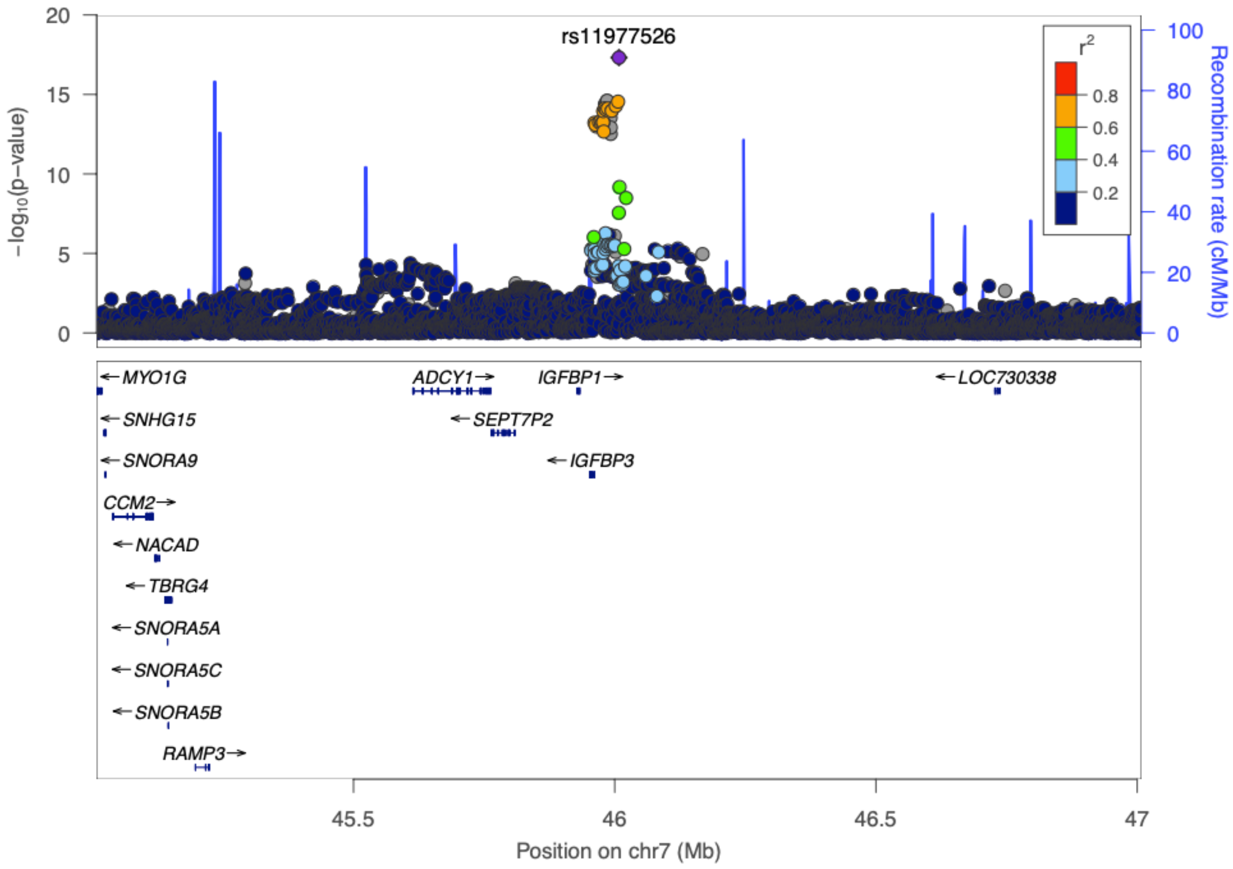

Chromosome 9 (near CDKN2B-AS1)

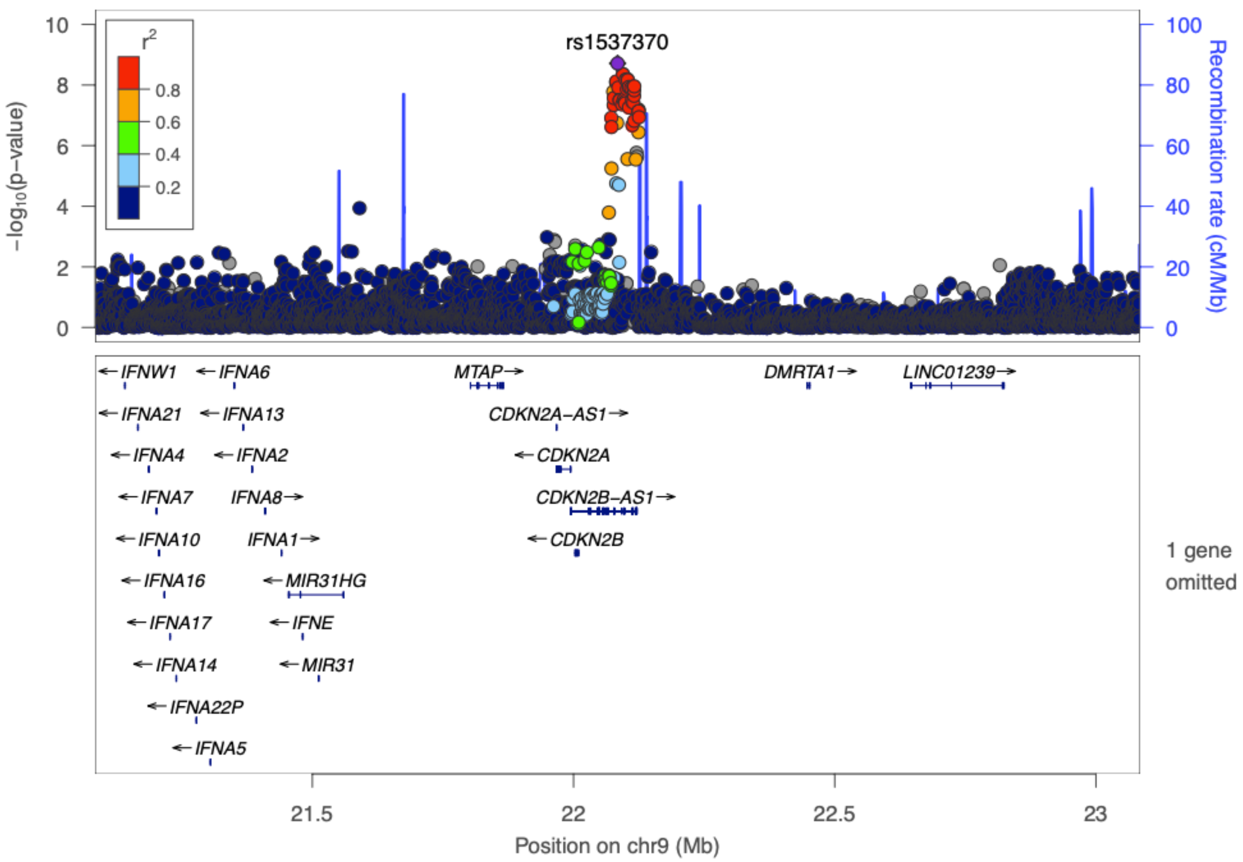

Chromosome 10 (near *CNNM2* & *NT5C2*)

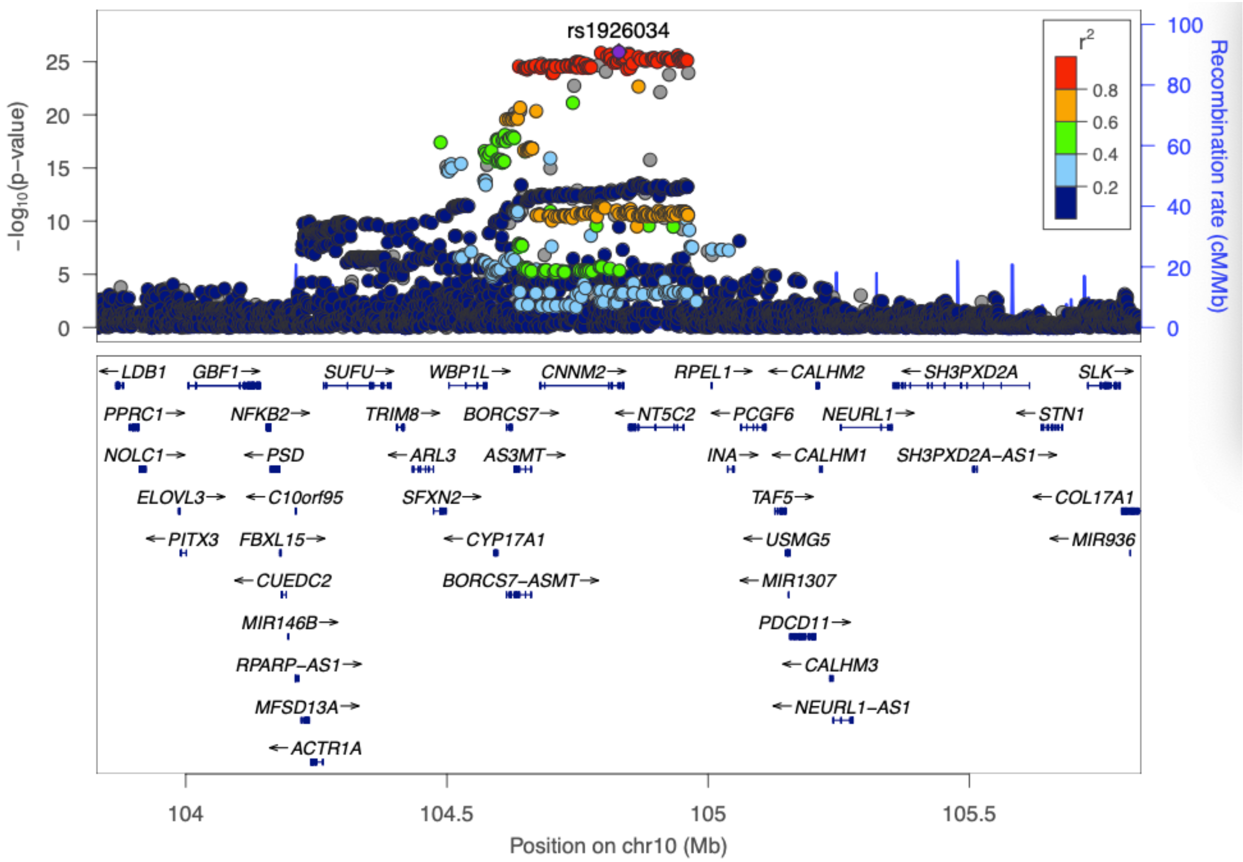

Chromosome 10 (near *AFAP1L2*)

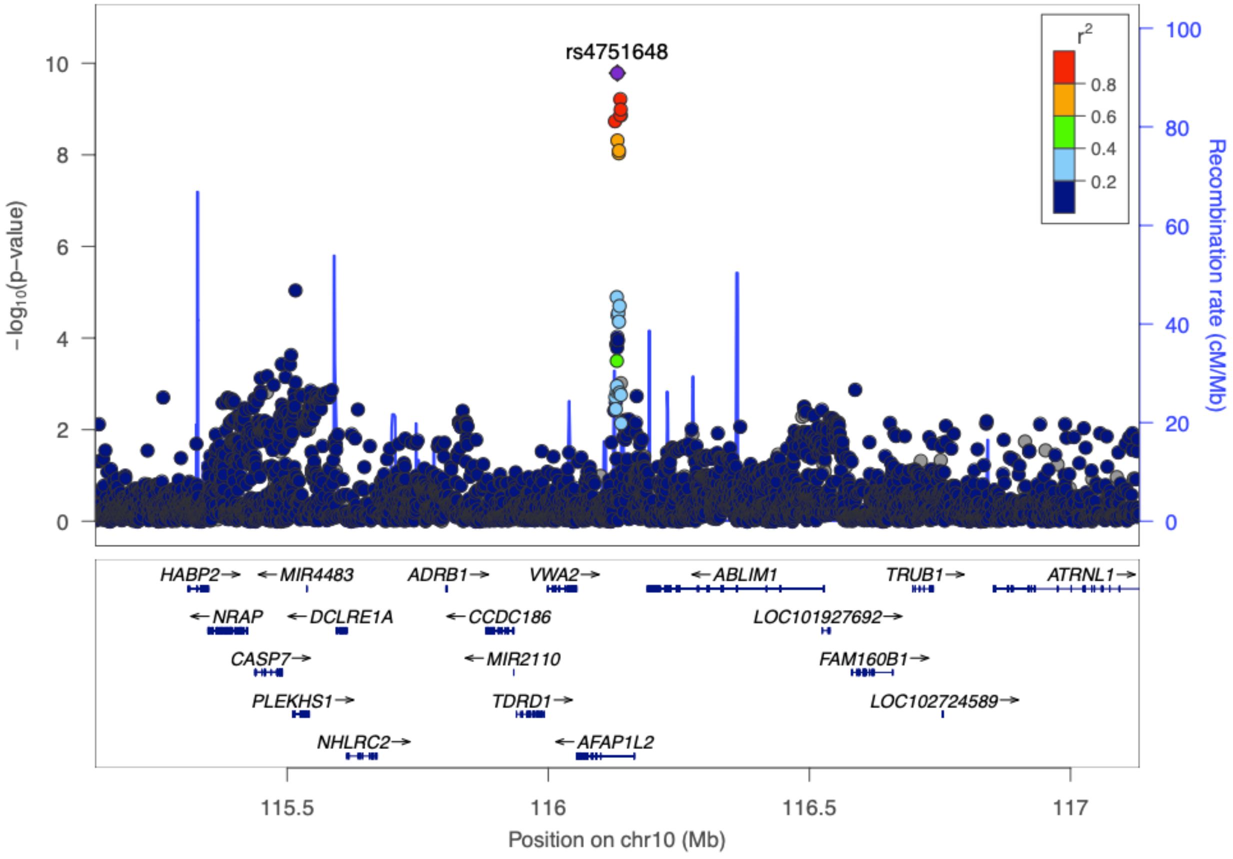

Chromosome 11 (near *MYBPC3*)

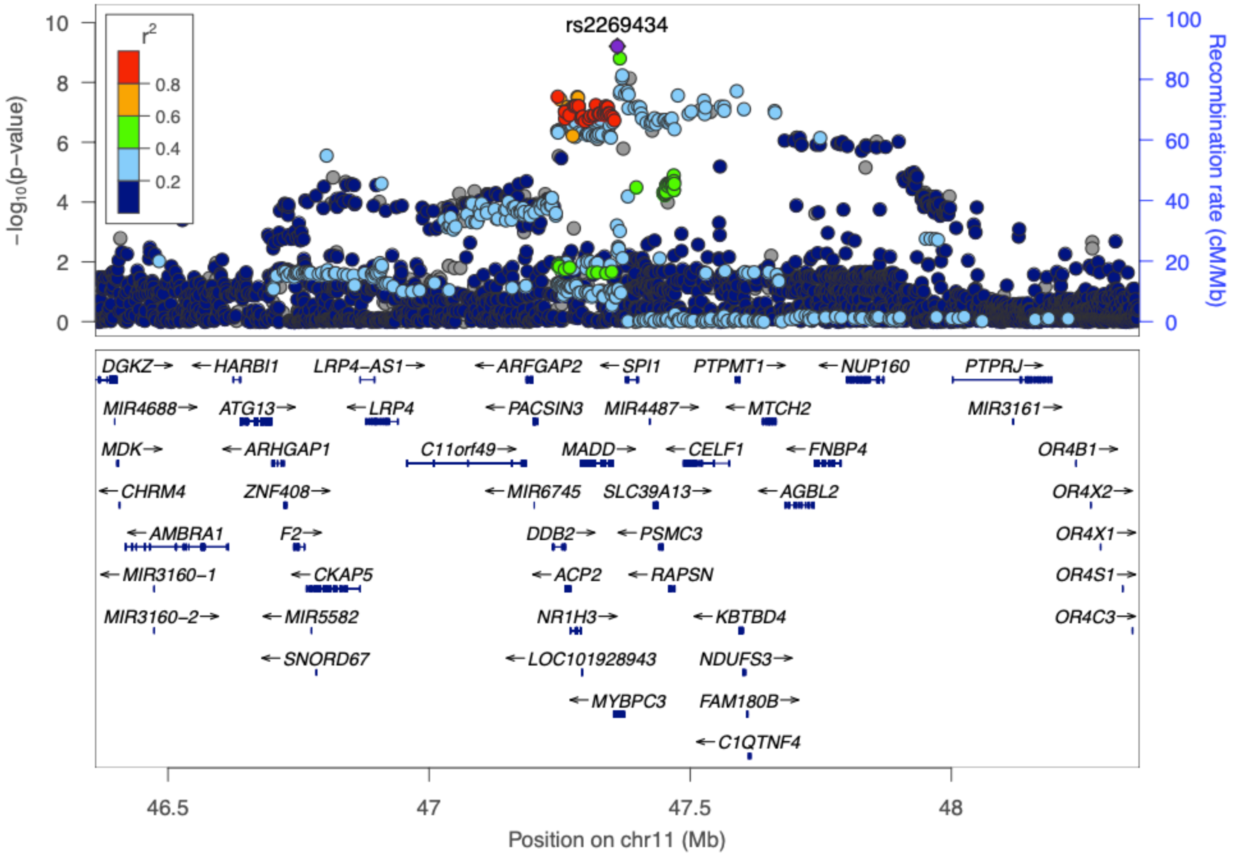

Chromosome 11 (near *ANO1*)

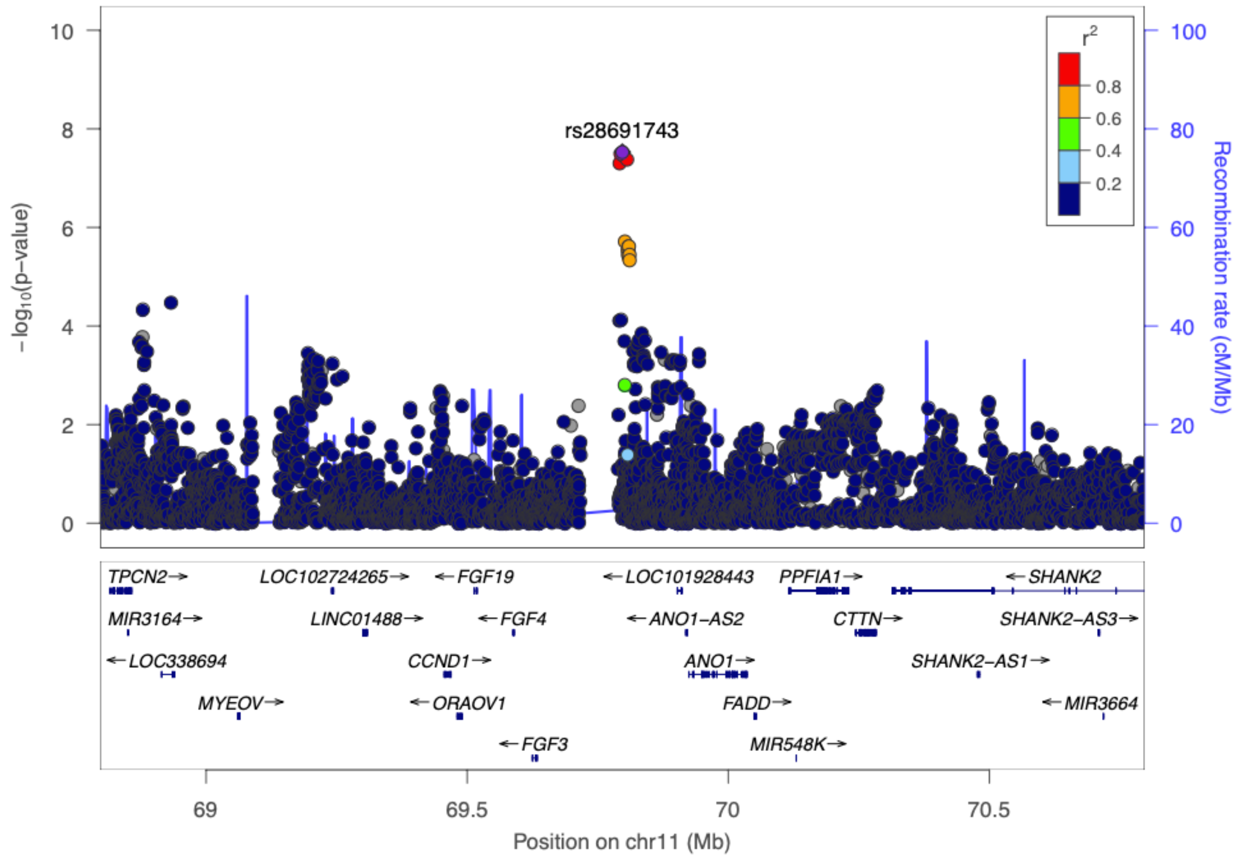

Chromosome 12 (near *LOC100506393*)

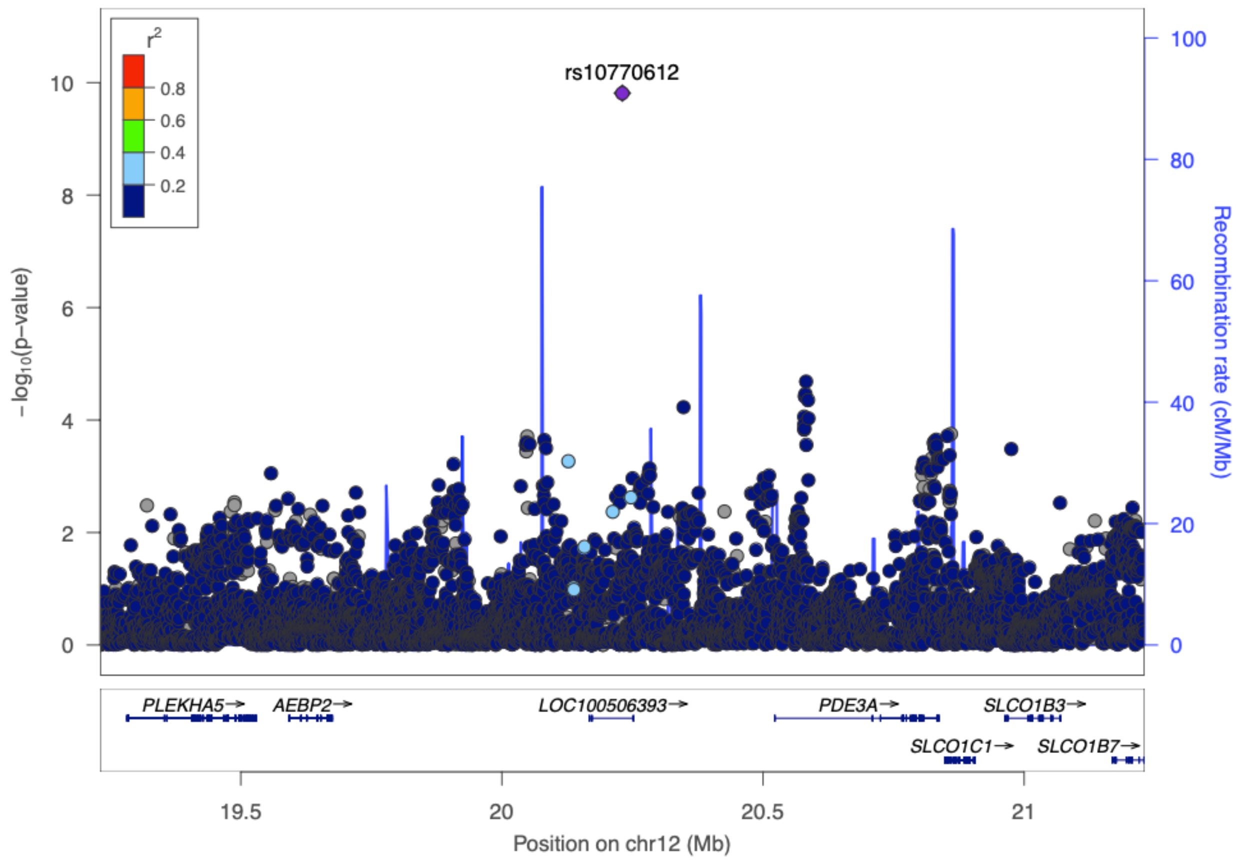

Chromosome 13 (near *COL4A2*)

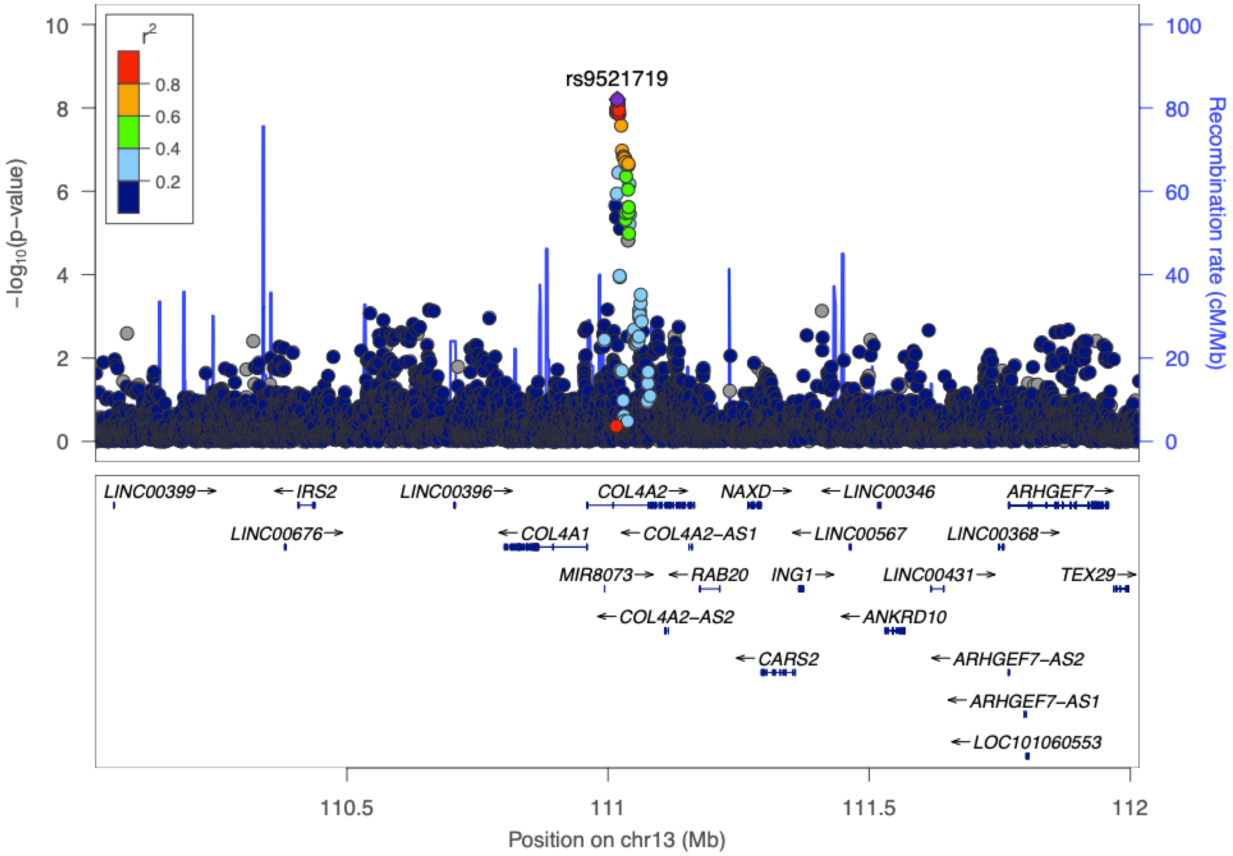

Chromosome 14 (near *SMOC1*)

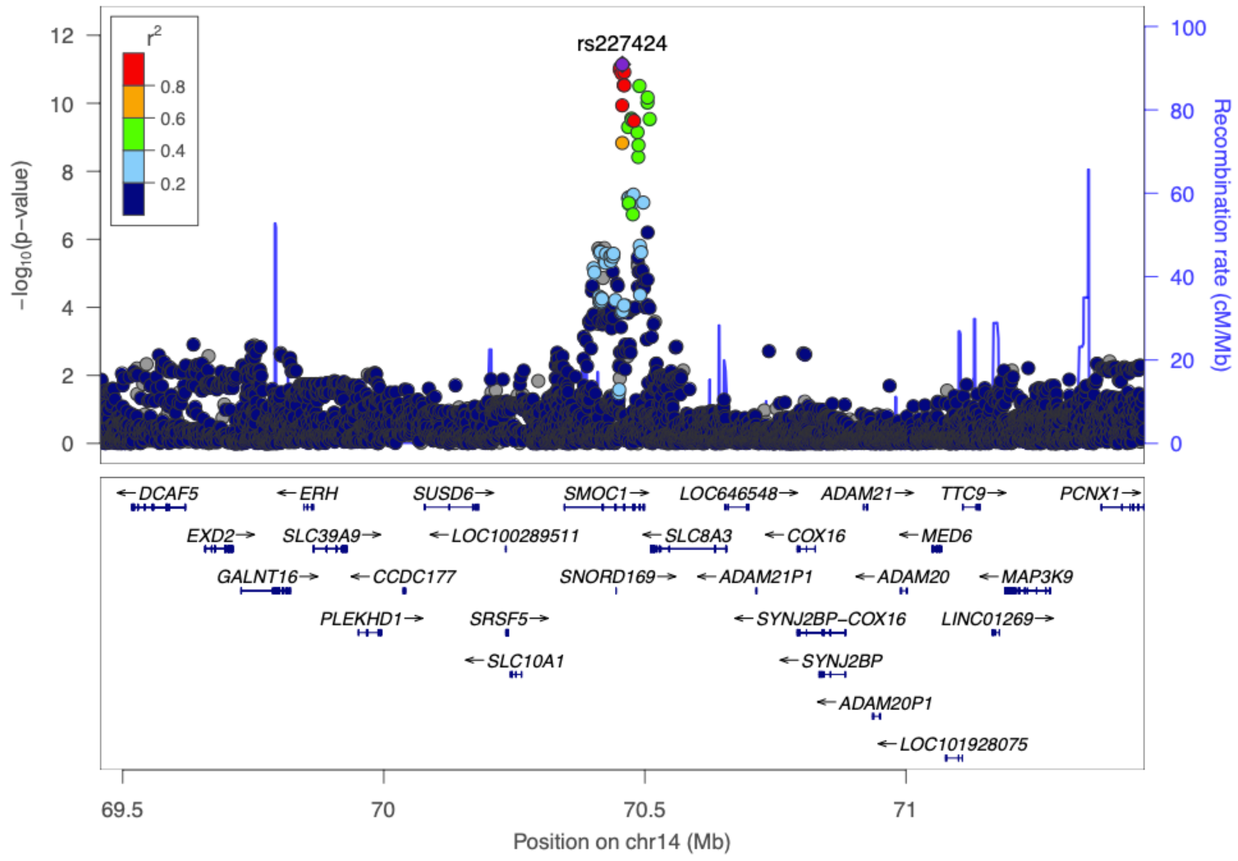

Chromosome 15 (near FURIN)

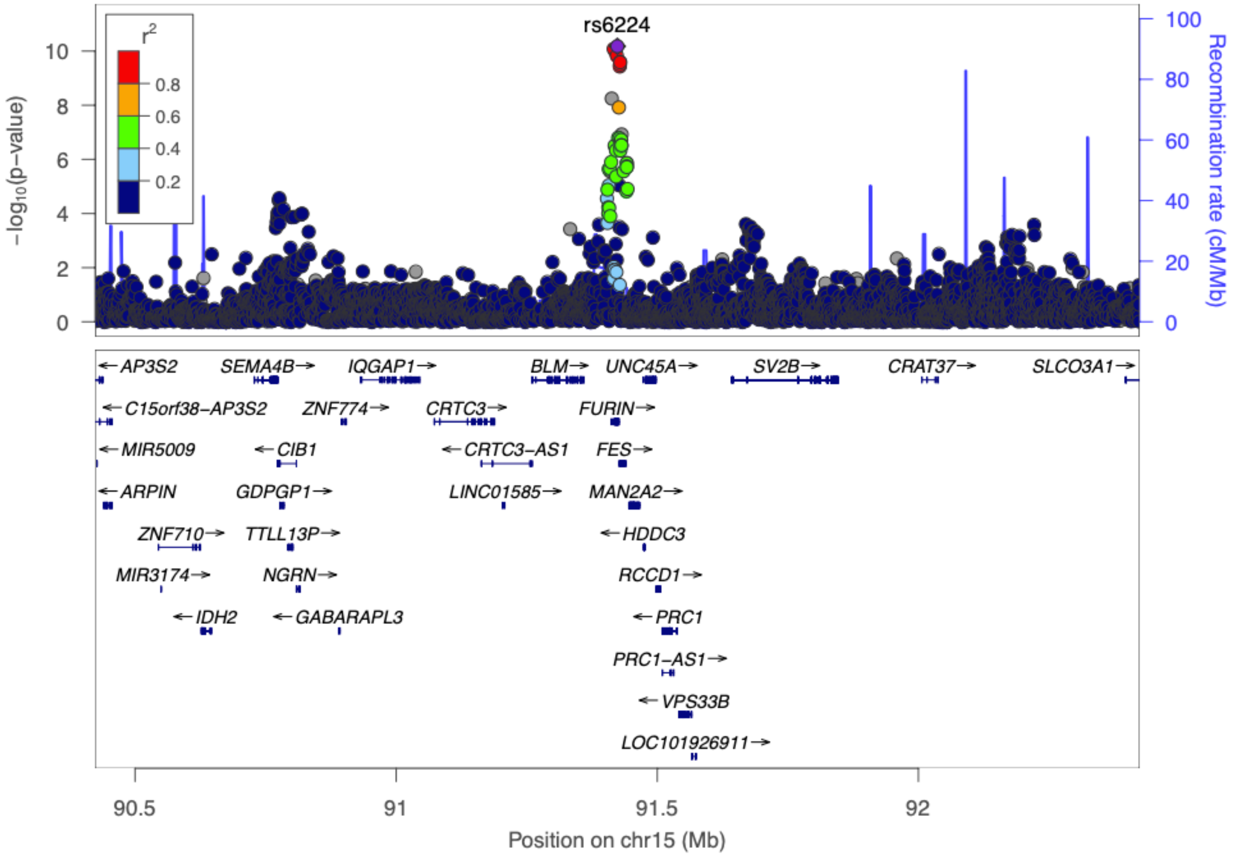
